# Recognizing the Evolution of Clinical Syndrome Spectrum Progression in Individuals with Single Large-Scale mitochondrial DNA deletion syndromes (SLSMDS)

**DOI:** 10.1101/2024.08.23.24312119

**Authors:** Rebecca Ganetzky, Katelynn D. Stanley, Laura E. MacMullen, Ibrahim George-Sankoh, Jing Wang, Amy Goldstein, Rui Xiao, Marni J. Falk

## Abstract

**Introduction:** Single large-scale mtDNA deletions (SLSMD) result in Single Large Scale Deletion Syndromes (SLSMDS). SLSMDS presentations have classically been recognized to encompass at least three distinct clinical phenotypes, Pearson Syndrome (PS), Kearns-Sayre Syndrome (KSS), and Chronic Progressive Ophthalmoplegia (CPEO).

**Methods:** Facilitated review of electronic medical records, manual charts, and REDCap research databases was performed to complete a retrospective natural history study of 32 SLSMDS participants in a single health system seen between 2002 and 2020. Characteristics evaluated included genetic and clinical laboratory test values, growth parameters, signs and symptoms, demographics, and patient reported outcome measures of fatigue, quality of life, and overall function.

**Results:** Detailed cohort characterization highlighted that a recurrent deleted region involving *MT-ND5* occurs in 96% of SLSMD subjects regardless of clinical phenotype, which tended to evolve over time. Higher blood heteroplasmy correlated with earlier age of onset. GDF-15 was elevated in all SLSMD subjects. A PS history yielded negative survival prognosis. Furthermore, increased fatigue and decreased quality of life were reported in SLSMD subjects with advancing age.

**Conclusion:** Retrospective natural history study of SLSMDS subjects demonstrated the evolution of classically considered PS, KSS, and CPEO clinical presentations within affected individuals, which may inform future clinical trial development.

## INTRODUCTION

Primary mitochondrial disease (PMD) is the most common category of inborn errors of metabolism, with over 400 distinct genetic etiologies^1,2^. Despite sharing a common pathogenesis of impaired energy production, each PMD may contain extensive phenotypic diversity^3,4^. Describing the scope of diversity within major genetic etiologies of PMD is critical to enable future development of precision therapies and drug trials with appropriate outcome measures that are clinically meaningful for each patient.

Mitochondria contain mitochondrial DNA (mtDNA), which is a 16,569 base-pair circular genome that encodes critical components of the oxidative phosphorylation system necessary to generate cellular energy. Multiple mtDNA genome copies exist within each mitochondrion, where hundreds to thousands of mitochondria may exist per cell. When a pathogenic variant or copy number alteration occurs within the mtDNA genome, it may be heteroplasmic, present in only a proportion of the mtDNA molecules within a given cell, tissue, or organism. Single large-scale mtDNA deletions (SLSMD) have been causally associated with a phenotypically diverse range of Single Large-Scale Mitochondrial DNA Deletion Syndromes (SLSMDSs). The most common SLSMD is a 4,977 base-pair deletion that begins at m.8482 and stretches to m.13459^5^. However, SLSMDs may vary in size, length and heteroplasmy level^5^. While most SLSMDs arise de novo at the early oocyte stage^6^, SLSMDs may result from pathogenic variants in nuclear genes^7^ related to regulation of mtDNA dynamics.

SLSMDS presentations are clinically broad, encompassing a spectrum of at least three distinct phenotypes, namely Pearson Syndrome (PS), Kearns Sayre Syndrome (KSS), and Chronic Progressive Ophthalmoplegia (CPEO). Occasionally, Leigh syndrome spectrum may also be seen^7,8^. PS classically presents in infancy with transfusion-dependent sideroblastic anemia. Accompanying symptoms in PS may variably include multiple cytopenias, exocrine pancreatic insufficiency, distal renal tubular acidosis, lactic acidosis, and/or cirrhosis^9–12^. Children who survive the early phase of the disease may have resolution of bone marrow and pancreatic disease, appear to enter a quiescent period for several years, and then during mid-childhood develop progressive symptoms of KSS^13^.

Pearson syndrome is rare, with most knowledge arising from relatively small studies, making it challenging to determine complete and accurate natural history. However, survival has been reported to range from 20-60%, with the three largest studies reporting mortalities of 40% (2/5), 64% (7/11), and 22% (7/32)^9–11,14^. The interval timing of resolution of Pearson syndrome and clinical progression to KSS, the percentage of patients who go on to manifest as KSS, as well as how many KSS patients may have had subclinical prodromal symptoms is unknown. KSS is clinically defined as developing pigmentary retinopathy, CPEO, and one symptom among cerebellar ataxia, cardiac conduction abnormalities and/or elevated cerebrospinal fluid (CSF) protein concentration by age 20 years. KSS patients also often develop multiple endocrinopathies, renal glomerular disease, gastrointestinal dysmotility, and short stature and/or failure to thrive^8^. CPEO (or CPEO-plus (+) when extra-ocular symptoms are present) is defined by having CPEO, often accompanied by other signs common to KSS without meeting KSS criteria. Although PS, KSS and CPEO are the three named phenotypes in the SLSMDS spectrum, the spectrum is yet broader, and includes patients who do not meet any of these criteria^8,9^ (Here referred to as SLSMDS, not otherwise specified; SLSMDS-NOS).

To prepare for future clinical trials targeting SLSMDSs, we performed a retrospective electronic medical record (EMR) review of 30 SLSMDS participants and paper medical record review of 2 SLMDS participants at our center to systematically describe the molecular and clinical scope of their disease spectrum, as well as to identify common disease features and patient-reported outcome (PRO) measures that would be amenable to clinical trials.

## METHODS

### Cohort identification & classification

PMD participants were consented to Children’s Hospital of Philadelphia (CHOP) IRB Study #08-006177 (MJF, PI). Eligible decedents were also included. Participant Electronic medical record (EMR) data from years 2002 to 2020 was exported from the back-end data warehouse, Clarity, and combined with historic hematologic records from pre-2002, clinical research outcome measure data from a Research Electronic Data Capture (REDCap) database, using Alteryx version 2021.4.2.47844 for integration and modeling, and displayed for visualization and data analysis in Tableau (version 2021.4.5)^15–18^. Additional visualizations were created in RStudio (version 4.4.0)^19^. PMD participants were included in this retrospective data analysis study if they had molecular confirmation of a SLSMD. A tiered clinical definition was established to appropriately classify the SLMDS phenotypes. First, KSS clinical diagnosis was defined for SLSMDS patients in this cohort using established clinical criteria (onset of symptoms before age 20 years, pigmentary retinopathy, CPEO, and the presence of at least one of: cerebellar ataxia, cardiac conduction block and/or elevated CSF protein concentration). Second, SLSMDS patients who did not meet criteria for KSS were defined as PS if they had either transfusion-dependent bone marrow failure and/or pancreatic exocrine insufficiency.

Third, remaining SLSMDS patients who met none of those criteria were defined as CPEO+ if they had, at minimum, clinically documented CPEO. All subjects had extra-ocular symptoms present and were therefore classified as CPEO+. Finally, remaining SLSMDS participants who failed to meet any of the preceding criteria were defined as SLSMDS-NOS (Figure 1A, Table 1B).

**Figure 1.**
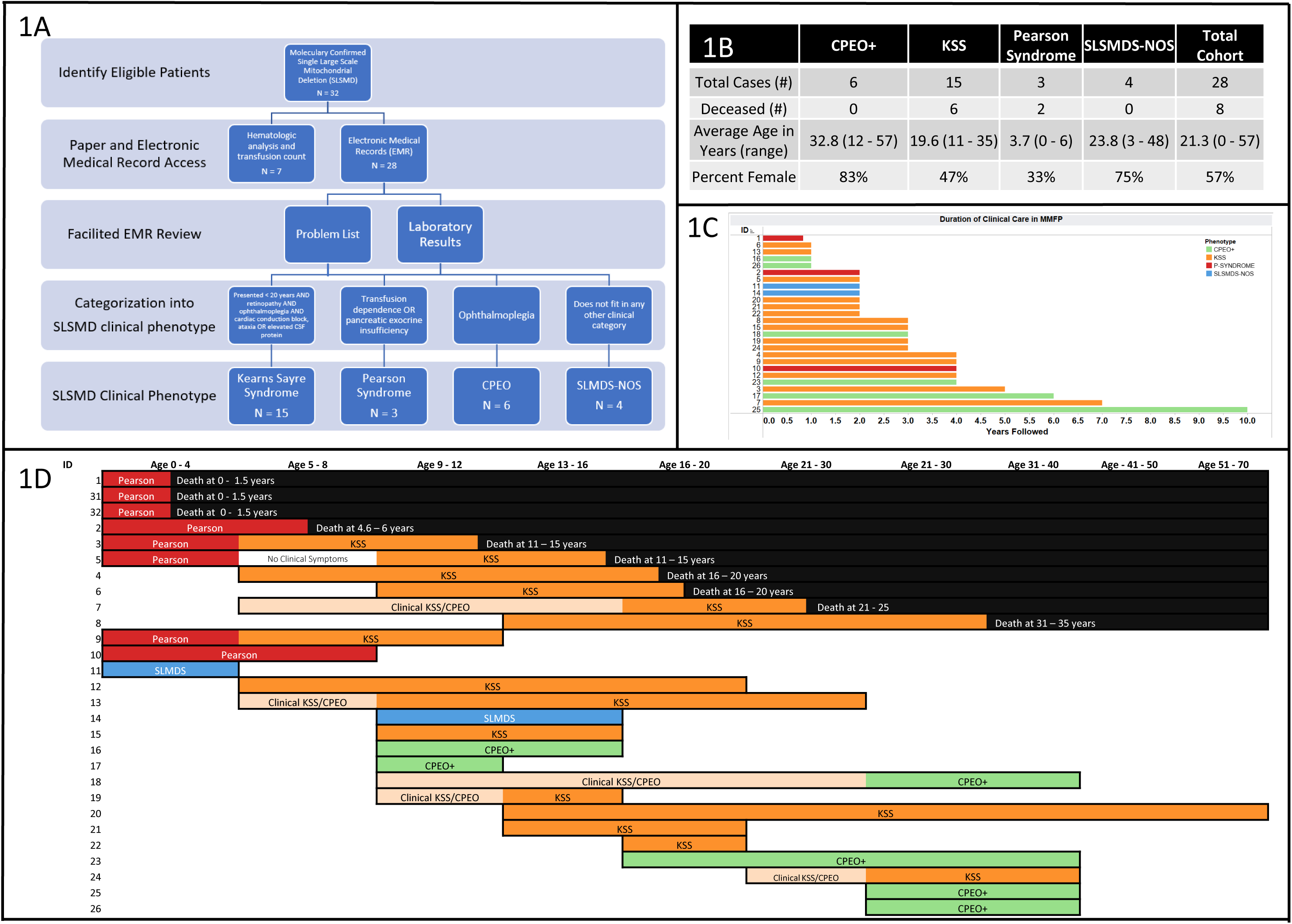
Description of clinical cohort. A. Inclusion and exclusion criteria and cohort definition flowchart. B. Table describing age distribution and living status of cohort. C Years Followed by MMFP. Colors (Red: Pearson Syndrome; Orange: KSS; Green: CPEO/CPEO+; Blue: SLSMDS-NOS) indicate the final phenotype for each participant. D. Timeline showing each patients clinical progression, including documented phenotypes prior to genetic confirmation. 2 participants SLSMDS-NOS, did not have data available regarding date of diagnosis. Includes 2 participant, 31 and 32, that did not have an EMR.

### Automated data review

Data elements extracted from the EMR (Epic Systems, Madison, WI) included physician-entered problem list, laboratory values, growth parameters, medications, age at first visit in the Mitochondrial Medicine Frontier Program (MMFP), demographic information (age, sex, race), and survival documentation.

### Facilitated Data Review

Three participants had clinical documentation that predated the initiation of the medical record system. Additionally, some data were missing from the automated data review. For the information that predated the EMR, paper medical charts were retrieved and manually reviewed for age of symptom onset, organ system involvement, genetic and laboratory testing results, and demographic information.

Missing data points were obtained through manual chart review. Data types that were not available as discrete fields in the EMR included genetic testing result details, and values of hematologic nadir, number of lifetime transfusions, and length of transfusion dependence.

### Growth Parameters

Z-scores for height, weight, and BMI were calculated using population means reported by the World Health Organization (WHO) for ages 2 and younger and by the U.S. Department of Health and Human Services Centers for Disease Control (CDC) for ages 2 to 20 using R packages ‘chldsds’ and ‘zscorer’^20–23^.

Visualization of growth trends across ages was done using smoothed conditional means implemented in R ggplot2 (version 3.5.0) function “geom_smooth”.

### Patient-Reported Outcome (PRO) measures

Patient reported outcome (PRO) measures were extracted from a dedicated REDCap database hosted at the CHOP. PROs were administered to participants since 2019 as a part of their clinical care in MMFP, sent through email and completed by either the caregiver or the participant themselves^14^.

Surveys were typically completed on an age-based interval, unless otherwise clinically indicated, with a frequency of 3-month intervals for ages 0 – 23 months, 6-month intervals for ages 2 – 17 years, and 12-month intervals for ages 18 and above.

Fatigue levels were measured through the Modified Fatigue Impact Scale (MFIS), a 21-question assessment on fatigue levels within the past 4 weeks of administration^24^. The scores for MFIS range from 0 to 84, with higher scores indicating higher levels of fatigue. Generally, scores above 38 are indicative of fatigue^25^.

Functional Status was measured through either the Karnofsky scale for ages 16 and older^26^ or the Lansky scale for ages 0 – 16^27^. These assessments consist of a single scale from 10 to 100. Lower scores indicate higher levels of functional impairment, with 10 representing complete disability and 100 indicating an individual is fully active. Scores between 10 and 40 indicate moderate to severe restrictions, between 50 and 70 mild to moderate restrictions, and 80 to 100 mild to no restrictions.

Health-related Quality of Life (QoL) was measured through the Pediatric Quality of Life Inventory (PedsQL) Generic Core Scales^28^ and PedsQL Infant Scales^29^. The specific scale administered was age dependent: Infant (ages 0-12 months), Infant (ages 13 – 23), Toddler (ages 2 – 4), Young Child (ages 5 to 7), Child (ages 8 – 12), Teen (ages 13 – 17), and Adult (ages 18 and above). The PedsQL evaluates health related quality of life across four dimensions: Physical Functioning, Emotional Functioning, Social Functioning, and School/Work Functioning (2 years of age and older) or Cognitive Functioning (infants aged 0-2 years old). These dimensions are then combined to report the total score. The total score for the PedsQL ranges from 0 to 100, with lower scores indicating lower self-reported or caregiver-reported quality of life. Raw total scores are presented, with the average normative score across all ages of 84.02 presented for reference^29–31^.

### Statistical Analyses

Survival curves were obtained by Kaplan-Meier method and survival differences between groups were assessed using log rank test implemented by the R packages “survival” and “survminer”. Differences between groups for continuous variables (deletion size) was calculated using Kruskal-Wallis testing because of the small sample size. The logarithmic correlation was used to assess relationship of age with the MFIS score and with the PedsQL score. Statistics were performed in R Studio version 2023.06.1+524 and Tableau version 2021.4.5.

## RESULTS

### Clinical Description of SLSMDS Participant Cohort

We identified 32 participants with molecularly-confirmed SLSMDS (Figure 1A). EMR data were available for facilitated review on 28 participants. Two participants who were only seen on a research basis were only included in genetic and demographic analyses. Two participants had medical records that predated the CHOP EMR and one participant had a combination of paper and CHOP EMR records available over the course of his life. For those three participants, hematologic data was collected from the paper chart, and the two on whom only paper data was available were excluded from further analysis. From the participant who had both EMR and paper data, a facilitated EMR review was performed. Of the analyzed 26 participants with electronic medical record data, 15 were clinically categorized as KSS, 3 as PS, 6 as CPEO and 2 as SLSMDS-NOS (Figure 1B). 2 of 16 (12.5%) KSS participants had previously met diagnostic criteria for PS (one had been diagnosed with PS; a second had a previously clinical diagnosis of “aplastic anemia”). A KSS participant with an SSBP1 *de novo* pathogenic variant was previously described^7^, as were five additional participants^32^.

Participants had been clinically followed for an average of 3.15 years (Figure 1C). The average age of the total SLSMDS cohort was 21.3 years (range 0 – 57). 8 SLSMDS participants were deceased by the time of analysis, including 2 PS and 6 KSS participants (one of whom had a preceding diagnosis of PS). The average age at time of death was 14.9 years (range 0 to 31). The cohort included 16 female and 12 male participants (Figure 1B). There was a broad age range of participants followed; in general, a trend to longer-length of follow-up was seen for older participants. PS participants and PS survivors were significantly younger than the rest of the cohort and were younger than age 6. As such PS participants age ranges were divided into quartiles (0 – 1.4 years, 1.6 – 3 years, 3.1 – 4.5 years, and 4.6 – 6 years). The rest of the cohort ages were divided in 5-year age ranges (0 – 5, 6 – 10, ect.). The clinical course of each SLSMDS participant in the cohort, including their clinical diagnoses and outcomes is depicted in Figure 1D.

### SLSMD Molecular Genetic Analysis

Clinical genetic diagnostic testing results that defined the size and location of the SLSMD was available for 24 participants. The recurrent ∼ 5 kilobase (Kb) “common deletion” was seen in 5/20 (25%) SLSMDS cases. The average SLSMD size in this cohort was 5.7 Kb (range 2.7 to 9.3; Figure 2A). No association was apparent between deletion size and clinical phenotype (p = 0.34), Figure 2B). The starting location of the deletion ranged from m.5245-12584, and the ending location from m.9908-16085. While no commonly deleted area was shared by all participants, 23/24 (95.8%) shared a deletion of the region from m.12584-m.13444, encoding a portion of *MT-ND5* (Figure 2C).

**Figure 2.**
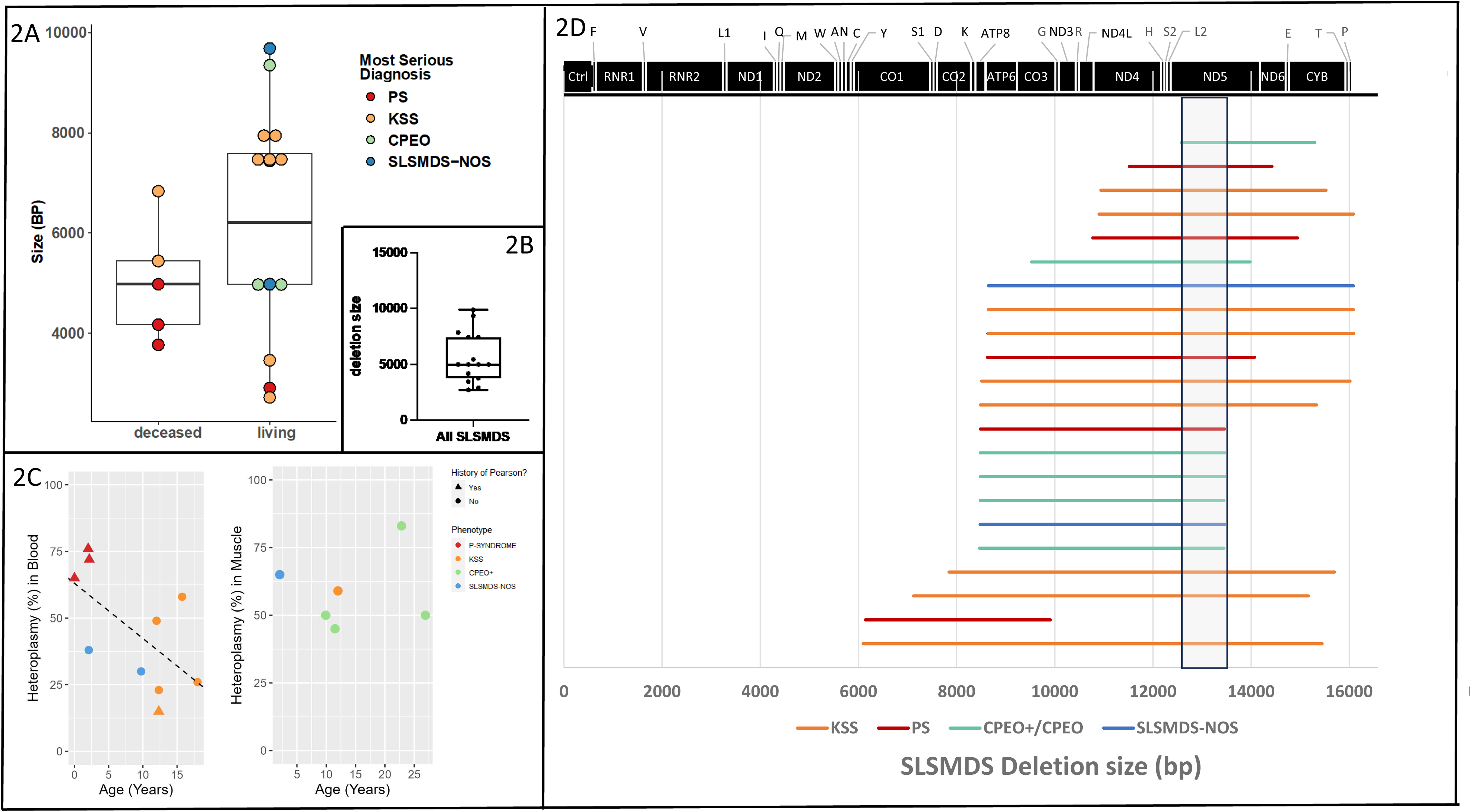
Genetic data. B. SLSMD deletion size in deceased (left) v. living participants (right) shows no correlation between deletion size and survival, or with most serious clinical phenotype (colors). Each bar represents a participant in the cohort, deletion data was not available on 6 participants in the cohort. B. A box plot of SLSMD deletion size in the cohort. C. Heteroplasmy by age at collection for both blood (left) and muscle (right). For the participant with 2 blood samples, their most recent test was used. Colors (Red = PS, Orange = KSS, Green = CPEO+, Blue = SLMDS-NOS) represent most recent phenotype classification. Triangles represent patients with a history of Pearson syndrome, while circles represent participants without a history of Pearson Syndrome. D. Depiction of each deletion location, colored by most severe SLSMDS clinical phenotype diagnosed during life span. Black line represents the mtDNA genome (depicted here as a line for visual convenience, starting on left from base pair position m.0). Box: shared deletion region

Testing methods used to quantify heteroplasmy levels varied, including quantitative PCR, digital droplet PCR, PCR followed by next generation sequencing and Southern Blot.

Heteroplasmy levels for the deletion may be overestimated by analytic methods that include polymerase chain reaction (PCR) amplification, which preferentially amplify the shorter (deleted mtDNA genome) amplicon^33^. SLSMD testing was performed in blood (n=19), buccal (n=2), saliva (n=3) and/or muscle (n=10) specimens. Of these, a numeric value for SLSMD heteroplasmy level (rather than a qualifier such as “large”) was provided for 11/19 blood, 3/3 buccal, 3/3 saliva, and 6/10 muscle assays. Average SLSMD heteroplasmy level across all available cases in blood was 45%, buccal 53%, saliva 17%, and muscle 59%. SLSMD heteroplasmy levels across all cases and sample types in the cohort ranged from 9-87%. Blood heteroplasmy levels appear to decline with age. PS participants had the highest heteroplasmy levels of the cohort. Muscle heteroplasmy also appears to decline with age. This analysis was limited by variations in the tissue studied and testing methodology performed (Figure 2C, Supplemental Table K).

Two participants had recurrent blood testing performed separated by a period of several years. In one case, the second heteroplasmy level, measured 3 years later appeared to increase from 25% to 58%, although the testing methods varied in these cases between the first and second assays. In the other case, the SLSMD was no longer detectable in blood on the repeat assay, which was approximately 5 years after the initial level taken in infancy, but was persistently detectable in muscle that was collected a month after the second blood sample. Interestingly, this second case was a PS survivor no longer transfusion dependent at the time of repeat deletion testing and was the only SLSMDS case in which the deletion was not detected in blood using NGS technology.

Four additional cases had SLSMD heteroplasmy levels clinically evaluated in more than one tissue type. One SLMDS participant had blood, muscle, and saliva heteroplasmy testing performed, their buccal (17%) and muscle (45%) was collected about a month apart and their salvia (19%) was collected 3 years later. Two SLSMDS participants had both blood and muscle heteroplasmy testing performed. One of the SLSMDS participants, their blood (49%) and muscle (59%) samples were collected on the same day. The other SLSMDS participant had their blood (38%) and their muscle (65%) collected one week apart. In another SLSMDS participant, deletion heteroplasmy had been measured on both buccal (56%) and saliva (23%) samples collected one year later.

### SLSMDS Cohort Clinical Phenotypes

8/28 (28%) SLSMDS participants were deceased by the time of retrospective study performance (Figure 3A). Mortality was particularly high in children who had a history of PS (4/5 (80%) were deceased). 4/11 KSS participants without a history of PS had also passed away (36%). Conversely, no mortality was seen in the CPEO cohort.

**Figure 3.**
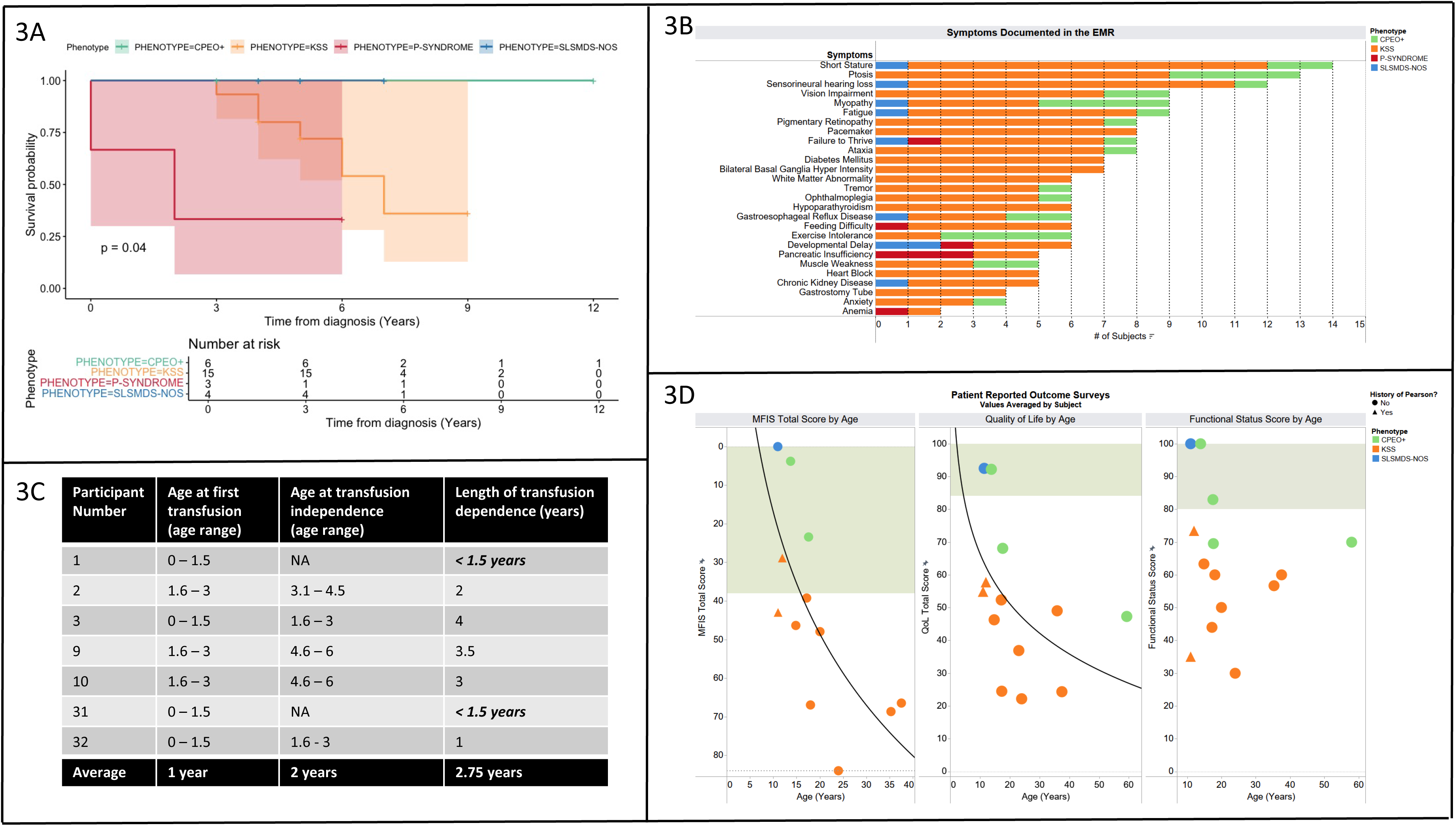
Clinical outcomes. A. Kaplan Meier curve of survival from the time of presentation, stratified by most severe phenotype (Red: Pearson Syndrome; Yellow: KSS; Green: CPEO/CPEO+; Blue: SLSMDS-NOS), with vertical lines representing censored data points. Table shows number of surviving participants by phenotype at each time point. There is a significant difference in survival from diagnosis by phenotype (p=0.04), with the lowest survival in Pearson syndrome, followed by KSS. No participants with CPEO/CPEO+ or SLSMDS-NOS were deceased in the time period followed. B. Rank list of documented symptoms by most current phenotype, sorted by frequency in all participants with SLSMDS. Data are color coded by participants most recent phenotype classification. Symptom must be present in at least 4 participants to be shown, apart from anemia, who had two participants documented. Data was abstracted from the medical record problem list and is not representative of all symptoms experience by the cohort. C. Table 1 Age at first transfusion and transfusion independence for patients with PS. Length of transfusion dependence was calculated as months between first and last transfusion; italicized cases (patient #A & D) passed away before achieving transfusion independence and were excluded from the average. Average age at first transfusion was 12 months; average age at transfusion independence was 48 months or 33 months after transfusion initiation. D. Z-scores for height by age. Colors (Red = PS, Orange = KSS, Green = CPEO+, Blue = SLMDS-NOS) represent most recent phenotype classification. Triangles represent patients with a history of Pearson syndrome, while circles represent participants without a history of Pearson Syndrome. Each line represents a different participant within the cohort, and unconnected lines indicate that the participant only had one datapoint. The black line is a smooth conditional means line to visualize average growth over time.

Clinical signs & symptoms were identified from the EMR “problem list” that were recurrent both within and across SLSMDS phenotypes. The most common problems seen in KSS were short stature (n=11), sensorineural hearing loss (n=10), ptosis (n=9), and need for pacemaker placement (n=8, including prophylactic placement for cardiac conduction abnormalities). Additional problems of note were vision impairment (n=7), fatigue (n=7), diabetes mellitus (n=7), and ataxia (n=7). In CPEO, ptosis (n=4), myopathy (n=4), and exercise intolerance (n=4) were recurrent problems. Anemia and pancreatic insufficiency were the only universal symptoms in PS. In SLSMDS-NOS, developmental delay was the most common problem.

Clinical symptoms reported in at least 4 participants were compared to identify areas of overlap between classical SLSMDS phenotype. Interestingly, failure to thrive was the only problem seen across participants with each of the named SLSMDS types. PS and KSS shared in common feeding difficulty and developmental delay. In addition, persistent pancreatic insufficiency, and anemia were seen not only in PS but also in KSS participants with a history of PS. CPEO+ and KSS shared in common symptoms of short stature, ptosis, sensorineural hearing loss, vision impairment, myopathy, fatigue, ataxia, tremor, gastroesophageal reflux disease, muscle weakness, exercise intolerance and anxiety. Perhaps surprisingly, no recurrent clinical problem was shared between PS and CPEO+ except failure to thrive (Figure 3B).

### SLSMDS Cohort Growth Assessment

Anthropometric data was available for 19 SLSMDS participants, and 16 participants had longitudinal data available. Height and length z-scores ranged from 0 to −7.25. All but three participants had most recent height z-scores at or below −2, indicating short stature. Longitudinally for height and length, KSS and PS participants grew negatively away from the growth curve. One CPEO+ participant grew in height positively with the growth curve and the other CPEO+ participant had an initial growth in height, before trending downwards again. Overall, as a cohort, growth was slow and short stature worsened with age with decreasing height z-scores over time; no participant experienced “catch-up” growth, including participants who were treated with recombinant human growth hormone (Supplemental Figure A).

BMI z-scores ranged from 1.75 to −10.20. Most participants trended at or above −2 z-score for BMI and longitudinally had growth parallel with the growth curve, but 3 KSS participants trended negatively along the BMI growth curve as they aged. 3 participants (2 KSS and 1 PS) had their most recent BMI z-score fall below −2, indicative of being underweight (Supplemental Figures C, D).

### SLSMDS Cohort PRO Results

Patient reported outcome measures were available for 13 SLSMDS participants, including one participant on whom all PRO assessments were completed except for MFIS fatigue scale. For participants with multiple scores from repeat assessments over time, fatigue scores and age at the time of completion were averaged participant. Individual longitudinal data is available in the appendix (Supplemental Figures H, J, I).

SLSMDS cohort average MFIS fatigue score was 42.35 (range 0 - 84), with higher numbers indicating greater levels of fatigue. 4 participants had values below the cut-off for fatigue, while 8 participants had scores above the normal range. One participant with KSS had the maximum score of 84 indicating high levels of fatigue, while another participant with SLSMDS-NOS had a score of zero indicating no reportable fatigue. While fatigue was a highly variable symptom across all SLSMDS participants as a whole, it was prevalent in nearly all individuals with KSS (7/8, 87.5%).

SLSMDS cohort average PedsQL score was 51.60 (range 19.44 – 92.15), with lower scores indicating lower self-reported or caregiver-reported health related quality of life. All but two participants fell below the normative score for their age category. Thus, QoL was commonly reduced below the normal range in the SLSMDS cohort, with QoL reduced in half (2/4, 50%) of the CPEO+ cohort and universally reduced in all KSS participants (9/9, 100%).

SLSMDS cohort average functional status score was 64.53 (range 30 – 100), with lower scores indicating lower levels of functional status. 53.33% of SLSMDS participants had self-reported or caregiver-reported scores showing mild to moderate restrictions in their functional status, with a higher proportion of KSS participants (60%) having mild to moderate restrictions compared to CPEO+ and SLMDS-NOS participants. Three participants, 2 CPEO+ and 1 SLMDS-NOS, had scores that were 80 or above, indicating minor restrictions, and two of these participants had scores of 100 indicating that they were fully active. No KSS participants had scores in this range. 4 SLSMDS participants, all with KSS had a scores below 50, indicating severe to moderate restrictions. However, functional status scores were consistently lower for KSS participants than for SLSMDS participants with either CPEO+ or SLSMDS-NOS.

Interestingly, both MFIS (R^2^ = 0.51, p = 0.0087) and PedsQL (R^2^ = 0.25, p = 0.082) scores in the SLSMDS participant cohort declined with age (Figure 3D), suggesting progressive symptoms.

### Laboratory values

Growth differentiation factor 15 (GDF-15) is a putative biomarker for mitochondrial disease, with different sensitivities and specificities for different classes of mitochondrial diseases^34^. GDF-15 was elevated in all SLSMDS participants above the upper limit of normal (750 pg/mL) and was higher in older participants (Figure 4A). There is a strong linear correlation with GDF-15 and age among CPEO+ participants and KSS participants without a history of Pearson (R^2^ = .95, p =.01). The average GDF-15 across the SLSMDS cohort was 3,729 pg/mL (range 885 – >6,000). In 5 participants, the values were increased above the reportable upper limit threshold of 6,000 (Figure 4A).

**Figure 4.**
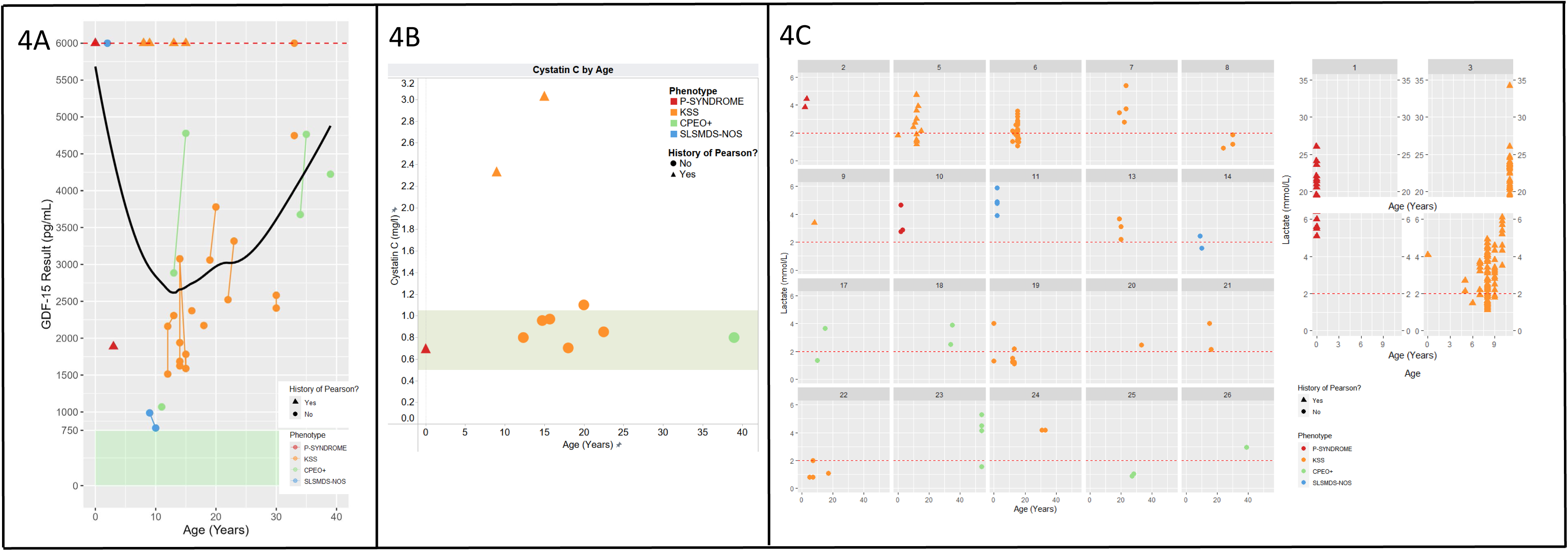
A. GDF-15 Levels by age and diagnosis. Each line represents a participant. Marks without connected lines indicates only one data point exists for that participant. The normal range (>750 pg/mL) is represented by a green box. All participants had abnormal values (>750 pg/mL), with one participant (SLSMDS) close to reference range. The dotted line (6000 pg/mL) is the maximum reportable value for this test, and 5 participants (1 PS, 1 SLSMDS-NOS, 3 KSS) had the maximum reportable range. Triangles represent patients with a history of Pearson syndrome, while circles represent patients without a history of Pearson Syndrome. Colors represent final clinical phenotype. The black line is a smoothed means line to visualize GDF-15 changes with age. B. Cystatin C levels by age and diagnosis. Values and ages are averaged by participant. Reference range (0.5 - 750 pg/mL) is highlighted in green. 2 participants with KSS had elevated Cystatin C. Triangles represent patients with a history of Pearson syndrome, while circles represent patients without a history of Pearson Syndrome. Colors represent final clinical phenotype. C. SLSMDS Cohort patient-reported outcomes by age and clinical syndrome diagnosis, PRO scales used are the Modified Impact Fatigue Scale (MFIS), the age appropriate PedsQL for Quality of Life (QoL), and the Karnofsky (age 16 or older) or Lansky Scale (15 or younger) for functional status. Scores are averaged by participant. The shaded areas of the graph represent normal ranges for the MFIS and QoL. For functional status, a score between 80 to 100 indicates minor to no functional impairment. Scores and ages are averaged by participant. Triangles represent patients with a history of Pearson syndrome, while circles represent patients without a history of Pearson Syndrome. No patients with active Pearson syndrome were surveyed. A correlation line was added for MFIS (x = 45.63*ln(y) + −88.19, R2 = 0.51, p = 0.0086) and PedsQL (x= −22.1039*ln(y) + 117.385, R2 = 0.25, p = 0.082) showing decline with age. D. Lactate levels for each participant by age show consistent elevation of lactate (left). Triangles represent patients with a history of Pearson syndrome, while circles represent patients without a history of Pearson Syndrome. Colors represent final clinical phenotype. Participants 1 and 3 have been plotted on a separate axis due to life-limiting severe lactic acidosis (right) with a break between 6 mmol/L and 20 mmol/L.

**Figure 5.**
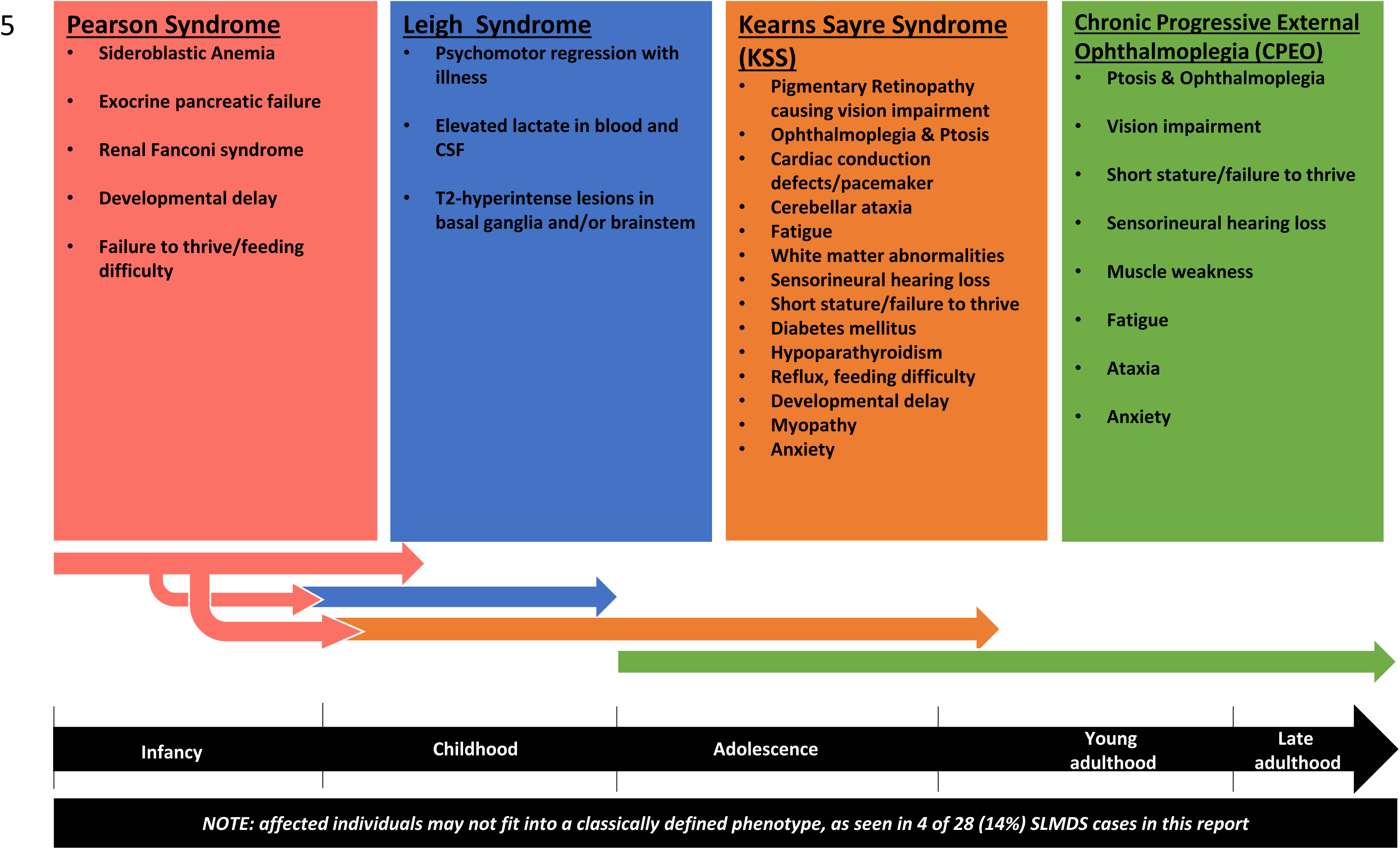
Spectrum of SLSMDS throughout the lifespan with common symptoms in this study emphasized when applicable (PS, KSS, CPEO). Each colored arrow corresponds to potential range of ages for symptom onset. Patients may transition between phenotypes over time, shown by branching arrows. In this study, 3/6 patients with PS developed KSS. Late adulthood represents > 30 years of age.

Cystatin C level is a marker of glomerular function that is independent of muscle bulk. Cystatin C values were available for 10 participants with an average value of 1.224 mg/L (range 0.7 – 3.03) and a median of 0.9. Two participants, both PS survivors, had significant glomerular dysfunction as evidenced by Cystatin C of 2.33 mg/L and 3.03 mg/L.

Lactate levels varied significantly both between and among individual participants. 17 of 24 (70%) participants had an elevated baseline lactate (>2 mmol/L), including all six participants with a history of PS (Figure 4C). There was not an overall correlation between lactate levels and outcomes, except for two participants with a history of PS who ultimately passed away from fatal lactic acidosis (Figure 4C, right panel).

### Hematologic manifestations of SLSMDS

Hematologic data was available on 7 participants with a history of PS, including 2 participants who were otherwise excluded from the study for limited EMR data. Anemia was participant seen, consistent with study inclusion criteria definition, in all PS participants. Anemia of renal disease was seen in two KSS participants with a history of PS and chronic kidney disease. These data points were excluded from analysis. No other participants had clinically significant anemia. The degree of anemia was more pronounced in participants who ultimately passed away (Figure 3C). All PS participants received at least one packed red blood cell (pRBC) transfusion (7/7, 100%), with an average of 27 lifetime pRBC transfusions (range 9-48) per participant.

Hematologic manifestations of SLSMDS in the PS cohort were analyzed in participants 1, 2, 3, 9 and 10. Participant 5 with a history of PS was excluded because no hematologic data were available. Participants 31 and 32, on whom only written medical records were available, were included for hematologic analysis, hospitalizations, and mortality.

Cytopenias extended beyond the red blood cell lineage. Interestingly, platelet transfusions were needed in 3/7 (42.8%) PS participants, with an average of 6 lifetime platelet transfusions (range 0-34). Neutropenia was seen in all PS participants, with an average absolute neutrophil count (ANC) nadir of 390 cells/uL (range 0-1,100, normal >1,500). Of note, one PS participant had a co-morbid clinical diagnosis of autosomal dominant benign cyclic neutropenia. All PS participants had multiple hospitalizations during their disease course, with an average of 10 lifetime hospital admissions (range 3-25). Transfusion independence was achieved by 5/7 (71%) patients, within an average of 2.8 years (range 1.5-4 years) after having received their first transfusion (Figure 3C).

Participant 1 died due to pseudomonal sepsis before transfusion independence; participants 3, 9 and 32 achieved transfusion independence but subsequently died. Both participants 3 and 9 died due to complications of stage V chronic kidney disease at ages 11 and 6, respectively. Participant 32 died of cardiac arrest 5 months after transfusion independence (no further data are available.) Participant 5 was not included in the transfusion data analysis but did have a history of transfusions per parental report and achieved transfusion independence prior to presentation at our center. He also ultimately died due to complications of stage V chronic kidney disease.

As mentioned above, anemia recurred in the two participants with KSS having a history of PS who ultimately died of stage 5 chronic kidney disease, which was considered to be anemia of renal disease. Subclinical anemia was also present in three additional KSS participants without a known PS history, one with proteinuria but without CKD, and two with CKD (stages 2 and 3). We postulate that the etiology of anemia in KSS may be multifactorial due to their chronic disease, and specifically may be anemia of renal disease. However, in participants with a history of PS it may also represent persistent cytopenia since in both cases it was more significant than in other KSS participants and predated clinical recognition of their renal disease; however, they also had more severe renal disease. No neutropenia or thrombocytopenia was seen outside of PS.

## DISCUSSION

Here, we performed a retrospective, single-center, multi-faceted, semi-automated clinical data longitudinal analysis in 28 SLSMDS participants, including in depth hematologic analysis on 2 additional PS participants on whom EMR data was not available. No correlation was seen of SLSMD deletion size or location with SLSMDS participants’ clinical phenotype and the “common” 5 Kb SLSMD was only present in 25% of cases.

However, we identified a 860 basepair deletion region extending from position m.12584-13444 that includes a portion of *MT-ND5* that was shared in common by 95.8% of all SLSMDS participants, an important finding that may help inform future studies into the pathogenetic mechanism of SLSMDS. SLSMD deletion size and heteroplasmy level analyses demonstrated great variability in all SLSMDS cases; however, there was a trend towards higher blood heteroplasmy correlating with earlier age of onset. As a spectrum disease, few phenotypic features were shared among all participants, but failure to thrive at some point throughout the life span was universal. The phenotype of SLSMDS participants evolved with age, with surviving participants from one presentation transitioning among other named clinical syndrome phenotypes, including from Pearson to KSS, and from “KSS/CPEO” overlap to CPEO+ in one case and from “KSS/CPEO” overlap to KSS in four cases. Our data suggest that some symptoms, such as fatigue, increase with age and quality of life declines. Although PS participants evolved to meet KSS criteria, they were distinct from other KSS participants by (1) having more significant anemia across the life span, which may be residual or related to kidney disease; (2) having younger age of onset of KSS features; and (3) having higher mortality than other KSS participants. Diagnostically, we demonstrated that GDF-15 appears to be a reliable disease biomarker of all SLSMDs, regardless of clinical phenotype. Although PRO assessments of clinically meaningful outcomes of fatigue, QoL, and functional status demonstrated variability among all SLSMDS participants, KSS participants consistently demonstrated high degrees of impairment, suggesting these PRO assessments should be considered as outcome measures in future clinical trials of candidate therapeutic interventions specifically for KSS.

SLSMDSs collectively are a common subset of PMD, presenting across all age ranges with a variably severe range of multi-system problems. The high morbidity and mortality of SLSMDSs highlights the critical need to develop targeted therapies. However, performance of successful clinical trials for candidate therapies will require selecting objective endpoints that are representative of the disease burden in this participant cohort and likely to respond to candidate treatment. To facilitate optimal endpoint selection, expanded understanding is needed of the natural history of SLSMDS. PRO assessments of fatigue, QoL, and overall functioning performed in the SLSMD cohort revealed several promising outcome metrics for future clinical trial consideration. While fatigue was a highly variable symptom across all SLSMDS participants as a combined cohort, it was prevalent in nearly all individuals with KSS (7/8, 87.5%). QoL was more commonly reduced below the normal range in the SLSMDS cohort, with QoL reduced in half (2/4, 50%) of the CPEO+ cohort and universally reduced in all KSS participants) (9/9, 100%). Functional status scores were highly variable across the SLSMDS population, but consistently lower for KSS participants than for SLSMDS participants categorized as either CPEO+ or SLSMDS-NOS. Thus, fatigue (MFIS), QoL (PedsQL), and/or functional status (Karnofsky-Lansky) PRO assessments in KSS participants consistently demonstrated high degrees of impairment in outcomes that are of high patient importance and plausible as potentially responsive metrics in future clinical trials of therapeutic interventions^4^.

Molecular genetic investigations of SLSMDS patients suggested that there are no genetic characteristics that reliably differentiate among PS, KSS and CPEO+ clinical syndromes. Deletions occurred throughout the mtDNA genome, and no association was identified in location of breakpoints and size of deletion. PS participants did have higher levels of heteroplasmy compared to other phenotypes, but sample size may skew the data However, these findings were limited by variability in the diagnostic testing methodology for deletion detection and heteroplasmy quantification, but are consistent with previously reported findings^9–11^. Yet unlike in older studies^35^, SLSMD was detectable in blood in almost all participants who had this testing performed in the study (with the exception of one PS survivor), perhaps showing improved sensitivity of next-generation sequencing methodologies. In three participants who had undergone saliva mtDNA testing, observed heteroplasmy levels were lower than other tissues: 9% to 36%, including one case where muscle heteroplasmy was also evaluated and found to be 30% higher. This is consistent with previous observation that mtDNA deletion heteroplasmy is usually higher in muscle than in blood and buccal samples^36^.

SLSMDS has classically been broken into discrete clinical entities. However, this retrospective cohort study highlights that broad overlap occurs between different clinical problems that occur in participants across the SLSMDS spectrum. Indeed, as we anticipated, overlapping phenotypic features were seen in KSS cohort participants with those of either PS or CPEO+. However, no overlap was seen between CPEO+ and PS, which appear to represent far extremes of the clinically heterogeneous spectrum continuum. Further, two participants classified as SLSMDS-NOS did not meet criteria for any of the classically described SLSMDS phenotypes and had no overlap in clinical features with each other. Universal among all phenotypes was a history of failure to thrive, although cross-sectional BMI data were not as revealing. Short stature was also common. These insights support the extensive multi-system nature of SLSMDS, with much more nuanced clinical disease manifestations possible and progression over time between the classical syndromic categories.

The stark contrast between the consistent presence of SLSMDs with overlapping locations in the mtDNA genome and the extensive phenotypic breadth is striking and remains unexplained. As we move towards genetic and cell-based therapies that target the underlying genetic defect to treat the root cause of disease^37,38^, this phenotypic variability proves challenging by prohibiting selection of a single universal clinical endpoint that will be meaningful for all SLSMDS participants. Therefore, clinical trials will need to be designed thoughtfully to accommodate specific outcomes that fall within the clinical breadth of the condition.

Investigations were performed of several laboratory markers of PMD. It is known that some biomarkers, such as GDF-15, are elevated in a genotype-specific manner in participants with PMD^39^. It has previously been reported that some participants with SLSMDSs may have elevated GDF-15^39^. To our knowledge, this is the largest study showing consistent elevations of GDF-15 across all SLSMDS participants. This intriguing finding suggests the presence of a common mechanistic pathway despite extensive phenotypic diversity. Also intriguing is that GDF-15 levels in these participants tended to positively correlate with age, which coincides with declines in fatigue scores and quality of life. One potential confounding variable is that renal function also correlates with increasing age, which may be a contributor to rising GDF-15 levels. These correlations are suggestive that GDF-15 may have utility as a secondary endpoint or companion biomarker in evaluating therapeutic effects of candidate treatments for SLSMDS.

Mitochondrial augmentation therapy (MAT) is a cell-based therapy that has shown initial promise in SLSMDS ^40^. In MAT, peripheral hematopoietic stem cells are harvested from the participant, augmented *ex vivo* with wild-type mitochondrial to reduce the heteroplasmy of the mtDNA deletion and then provided back as an allogenic hematopoietic stem cell transplant. However, the penetrance and persistence of mitochondrial-enriched bone marrow stem cells into non-bone marrow sites is unknown, including most critically in the central nervous system. Understanding the progressive timing of symptom onset and prognostic indicators is critical to evaluate the potential efficacy of proposed cell, gene, or drug therapies for SLSMDSs.

Limitations of this study include the retrospective collection of data, with inherent variation in laboratory methods and sample types used for clinical diagnostic testing over time, as well as likely clinician differences in entering problems into the EMR problem list. Small sample sizes also precluded definitive interpretation of statistically significant differences between SLSMDS groups. However, this study demonstrates the feasibility of performing facilitated, semi-automated EMR and clinical outcomes research database reviews to gather and analyze longitudinal data on rare disease participants. As this approach is more efficient and lower-cost than conducting a longitudinal natural history study over decades, retrospective studies remain valuable tools to facilitate clinical trial readiness as genetic, cell-based, and novel drug therapies rapidly evolve for evaluation in rare genetic diseases.

## Data Availability

All data produced in the present work are contained in the manuscript

## ACKNOWLEDGEMENTS.

We are grateful to John Campbell and Bethany Sensenig at Minovia Theraeputics for helpful discussions on SLSMDS outcome considerations. This work was supported in part by an investigator-initiated sponsored research award from Minovia Therapeutics (Falk, PI) and from the Children’s Hospital of Philadelphia Mitochondrial Medicine Frontier Program.

## AUTHOR CONTRIBUTIONS

MJF designed the study and obtained project funding. IGS performed EMR data curation and integration. LM, KS, and RG generated data visualizations. AG, RG, and MJF participated in project direction and data interpretation. RG, KS, and IGS wrote the manuscript draft. RX provided statistical support All authors contributed to manuscript revisions and final approval.

## CONFLICTS OF INTEREST

RG is a consultant for Minovia Therapeutics and Nurture Genomics.

MJF is engaged with several companies involved in mitochondrial disease therapeutic preclinical and/or clinical-stage development. MJF is co-founder and Chief Scientific Advisor of Rarefy Therapeutics LLC; an advisory board member with equity interest in RiboNova Inc.; a scientific advisory board member and paid consultant with Khondrion, and Larimar Therapeutics; has served as a paid consultant for Astellas (formerly MitoBridge), Casma Therapeutics, Cyclerion Therapeutics, Imel Therapeutics, Mayflower, Inc., Minovia Therapeutics, Mission Therapeutics, Myto Therapeutics, NeuroVive Pharmaceutical AB, Precision Biosciences, Primera Therapeutics, Inc., Reneo Therapeutics, Stealth BioTherapeutics, and Vincere Bio; and/or a sponsored research collaborator for Adjuvia Therapeutics, Astellas, Cyclerion Therapeutics, Epirium Bio, Imel Therapeutics, Khondrion, Merck, Minovia Therapeutics, Mission Therapeutics, NeuroVive Pharmaceutical AB, PTC Therapeutics, Reneo Therapeutics, RiboNova, Saol Therapeutics, Standigm, and Stealth BioTherapeutics. MJF also has received royalties from Elsevier and speaker fees from Agios Pharmaceuticals and GenoMind.

AG is a paid consultant for Reneo Therapeutics, Cyclerion/Tisento Therapeutics, and UCB Therapeutics. None of the other authors have relevant conflicts of interest to declare.

## Data Availability

Data that support the findings of this study are available by individual request.

## Ethics Statement

All human participants research was performed per Children’s Hospital of Philadelphia (CHOP) Institutional Review Board approved study #08-6177 (Falk, PI). Informed consent was obtained from all participants as required by the Children’s Hospital of Philadelphia (CHOP) Institutional Review Board.

## Notes

### Author Declarations

All human participants research was performed per Childrens Hospital of Philadelphia (CHOP) Institutional Review Board approved study #08-6177 (Falk PI).

## REFERENCES

1. Gorman GS, Chinnery PF, DiMauro S, et al. Mitochondrial diseases. Nature reviews Disease primers. 2016;2:16080. doi:10.1038/nrdp.2016.80

2. McCormick EM, Muraresku CC, Falk MJ. Mitochondrial Genomics: A complex field now coming of age. Current genetic medicine reports. 2018;6(2):52–61. doi:10.1007/s40142-018-0137-x

3. Falk MJ. Mitochondrial Disease Genes Compendium: From Genes to Clinical Manifestations. (Falk MJ, ed.). Elsevier Academic Press; 2020.

4. Zolkipli-Cunningham Z, Xiao R, Stoddart A, et al. Mitochondrial disease patient motivations and barriers to participate in clinical trials. Reddy H, ed. PLOS ONE. 2018;13(5):e0197513. doi:10.1371/journal.pone.0197513

5. Mancuso M, Orsucci D, Angelini C, et al. Redefining phenotypes associated with mitochondrial DNA single deletion. J Neurol. 2015;262(5):1301–1309. doi:10.1007/s00415-015-7710-y

6. Chinnery PF, DiMauro S, Shanske S, et al. Risk of developing a mitochondrial DNA deletion disorder. Lancet. 2004;364(9434):592-596. doi:10.1016/S0140-6736(04)16851-7

7. Gustafson MA, McCormick EM, Perera L, et al. Mitochondrial single-stranded DNA binding protein novel de novo SSBP1 mutation in a child with single large-scale mtDNA deletion (SLSMD) clinically manifesting as Pearson, Kearns-Sayre, and Leigh syndromes. Mishmar D, ed. PLoS ONE. 2019;14(9):e0221829. doi:10.1371/journal.pone.0221829

8. Goldstein A, Falk MJ. Mitochondrial DNA Deletion Syndromes. (Pagon RA, Adam MP, Ardinger HH, et al., ed.). University of Washington, Seattle; 1993. Accessed May 11, 2022. http://www.ncbi.nlm.nih.gov/pubmed/20301382

9. Broomfield A, Sweeney MG, Woodward CE, et al. Paediatric single mitochondrial DNA deletion disorders: an overlapping spectrum of disease. Journal of inherited metabolic disease. 2015;38(3):445–457. doi:10.1007/s10545-014-9778-4

10. Anteneová N, Kelifová S, Kolářová H, et al. The Phenotypic Spectrum of 47 Czech Patients with Single, Large-Scale Mitochondrial DNA Deletions. Brain Sci. 2020;10(11):766. doi:10.3390/brainsci10110766

11. Farruggia P, Di Cataldo A, Pinto RM, et al. Pearson Syndrome: A Retrospective Cohort Study from the Marrow Failure Study Group of A.I.E.O.P. (Associazione Italiana Emato-Oncologia Pediatrica). JIMD reports. 2016;26:37–43. doi:10.1007/8904_2015_470

12. Yoshimi A, Ishikawa K, Niemeyer C, Grünert SC. Pearson syndrome: a multisystem mitochondrial disease with bone marrow failure. Orphanet J Rare Dis. 2022;17:379. doi:10.1186/s13023-022-02538-9

13. Reynolds E, Byrne M, Ganetzky R, Parikh S. Pediatric single large-scale mtDNA deletion syndromes: The power of patient reported outcomes. Molecular Genetics and Metabolism. 2021;134(4):301–308. doi:10.1016/j.ymgme.2021.11.004

14. Björkman K, Vissing J, Østergaard E, et al. Phenotypic spectrum and clinical course of single large-scale mitochondrial DNA deletion disease in the paediatric population: a multicentre study. J Med Genet. 2023;60(1):65–73. doi:10.1136/jmedgenet-2021-108006

15. MacMullen LE, George-Sankoh I, Stanley K, et al. Bridging the clinical-research gap: Harnessing an electronic data capture, integration, and visualization platform to systematically assess prospective patient-reported outcomes in mitochondrial medicine. Mol Genet Metab. 2024;142(1):108348. doi:10.1016/j.ymgme.2024.108348

16. George-Sankoh I, MacMullen LE, Chinwalla AT, et al. MMFP-Tableau: Enabling Precision Mitochondrial Medicine through Integration, Visualization, and Analytics of Clinical and Research Health System Electronic Data. medRxiv. Published online January 1, 2024:2024.01.03.24300791. doi:10.1101/2024.01.03.24300791

17. Harris PA, Taylor R, Minor BL, et al. The REDCap consortium: Building an international community of software platform partners. J Biomed Inform. 2019;95:103208. doi:10.1016/j.jbi.2019.103208

18. Harris PA, Taylor R, Thielke R, Payne J, Gonzalez N, Conde JG. Research electronic data capture (REDCap)--a metadata-driven methodology and workflow process for providing translational research informatics support. J Biomed Inform. 2009;42(2):377–381. doi:10.1016/j.jbi.2008.08.010

19. R Core Team. R: A Language and Environment for Statistical Computing. Published online 2024. https://www.R-project.org/

20. Organization WH. WHO Child Growth Standards : Growth Velocity Based on Weight, Length and Head Circumference : Methods and Development. World Health Organization; 2009. Accessed August 8, 2024. https://iris.who.int/handle/10665/44026

21. Fryar CD, Kruszon-Moran D, Gu Q, Carroll M, Ogden CL. Mean Body Weight, Height, Waist Circumference, and Body Mass Index Among Children and Adolescents: United States, 1999-2018. Natl Health Stat Report. 2021;(160):1–24.

22. Myatt M, Guevarra E. zscorer: Child Anthropometry z-Score Calculator. https://CRAN.R-project.org/package=zscorer

23. Vogel M. childsds: Data and Methods Around Reference Values in Pediatrics. https://CRAN.R-project.org/package=childsds

24. Larson RD. Psychometric properties of the modified fatigue impact scale. Int J MS Care. 2013;15(1):15–20. doi:10.7224/1537-2073.2012-019

25. Flachenecker P, Kümpfel T, Kallmann B, et al. Fatigue in multiple sclerosis: a comparison of different rating scales and correlation to clinical parameters. Mult Scler. 2002;8(6):523–526. doi:10.1191/1352458502ms839oa

26. Schag CC, Heinrich RL, Ganz PA. Karnofsky performance status revisited: reliability, validity, and guidelines. J Clin Oncol. 1984;2(3):187–193. doi:10.1200/JCO.1984.2.3.187

27. Lansky SB, List MA, Lansky LL, Ritter-Sterr C, Miller DR. The measurement of performance in childhood cancer patients. Cancer. 1987;60(7):1651–1656. doi:10.1002/1097-0142(19871001)60:7<1651::aid-cncr2820600738>3.0.co;2-j

28. Varni JW, Seid M, Kurtin PS. PedsQL 4.0: reliability and validity of the Pediatric Quality of Life Inventory version 4.0 generic core scales in healthy and patient populations. Med Care. 2001;39(8):800–812. doi:10.1097/00005650-200108000-00006

29. Varni JW, Limbers CA, Neighbors K, et al. The PedsQL^TM^ Infant Scales: feasibility, internal consistency reliability, and validity in healthy and ill infants. Qual Life Res. 2011;20(1):45–55. doi:10.1007/s11136-010-9730-5

30. Varni JW, Burwinkle TM, Seid M, Skarr D. The PedsQL 4.0 as a pediatric population health measure: feasibility, reliability, and validity. Ambul Pediatr. 2003;3(6):329–341. doi:10.1367/1539-4409(2003)003<0329:tpaapp>2.0.co;2

31. Limperg PF, Haverman L, van Oers HA, van Rossum MAJ, Maurice-Stam H, Grootenhuis MA. Health related quality of life in Dutch young adults: psychometric properties of the PedsQL generic core scales young adult version. Health Qual Life Outcomes. 2014;12:9. doi:10.1186/1477-7525-12-9

32. Wild KT, Goldstein AC, Muraresku C, Ganetzky RD. Broadening the phenotypic spectrum of Pearson Syndrome: Five new cases and a review of the literature. Am J Med Genet A. 2020;182(2):365–373. doi:10.1002/ajmg.a.61433

33. Wang J, Balciuniene J, Diaz-Miranda MA, et al. Advanced approach for comprehensive mtDNA genome testing in mitochondrial disease. Mol Genet Metab. 2022;135(1):93–101. doi:10.1016/j.ymgme.2021.12.006

34. Yatsuga S, Fujita Y, Ishii A, et al. Growth differentiation factor 15 as a useful biomarker for mitochondrial disorders. Ann Neurol. 2015;78(5):814–823. doi:10.1002/ana.24506

35. Kleinle S, Wiesmann U, Superti-Furga A, et al. Detection and characterization of mitochondrial DNA rearrangements in Pearson and Kearns-Sayre syndromes by long PCR. Hum Genet. 1997;100(5-6):643–650. doi:10.1007/s004390050567

36. Jeppesen TD, Duno M, Vissing J. Mutation Load of Single, Large-Scale Deletions of mtDNA in Mitotic and Postmitotic Tissues. Front Genet. 2020;11:547638. doi:10.3389/fgene.2020.547638

37. Bueren JA, Auricchio A. Advances and Challenges in the Development of Gene Therapy Medicinal Products for Rare Diseases. Hum Gene Ther. 2023;34(17-18):763–775. doi:10.1089/hum.2023.152

38. Chancellor D, Barrett D, Nguyen-Jatkoe L, Millington S, Eckhardt F. The state of cell and gene therapy in 2023. Mol Ther. 2023;31(12):3376–3388. doi:10.1016/j.ymthe.2023.11.001

39. Lehtonen JM, Auranen M, Darin N, et al. Diagnostic value of serum biomarkers FGF21 and GDF15 compared to muscle sample in mitochondrial disease. Journal of Inherited Metabolic Disease. 2021;44(2):469–480. doi:10.1002/jimd.12307

40. Jacoby E, Bar-Yosef O, Gruber N, et al. Mitochondrial augmentation of hematopoietic stem cells in children with single large-scale mitochondrial DNA deletion syndromes. Sci Transl Med. 2022;14(676):eabo3724. doi:10.1126/scitranslmed.abo3724

