## Supplemental Figures for "Recognizing the Evolution of Clinical Syndrome Spectrum Progression in Individuals with Single Large-Scale mitochondrial DNA deletion syndromes (SLSMDS)"

### Slide 1
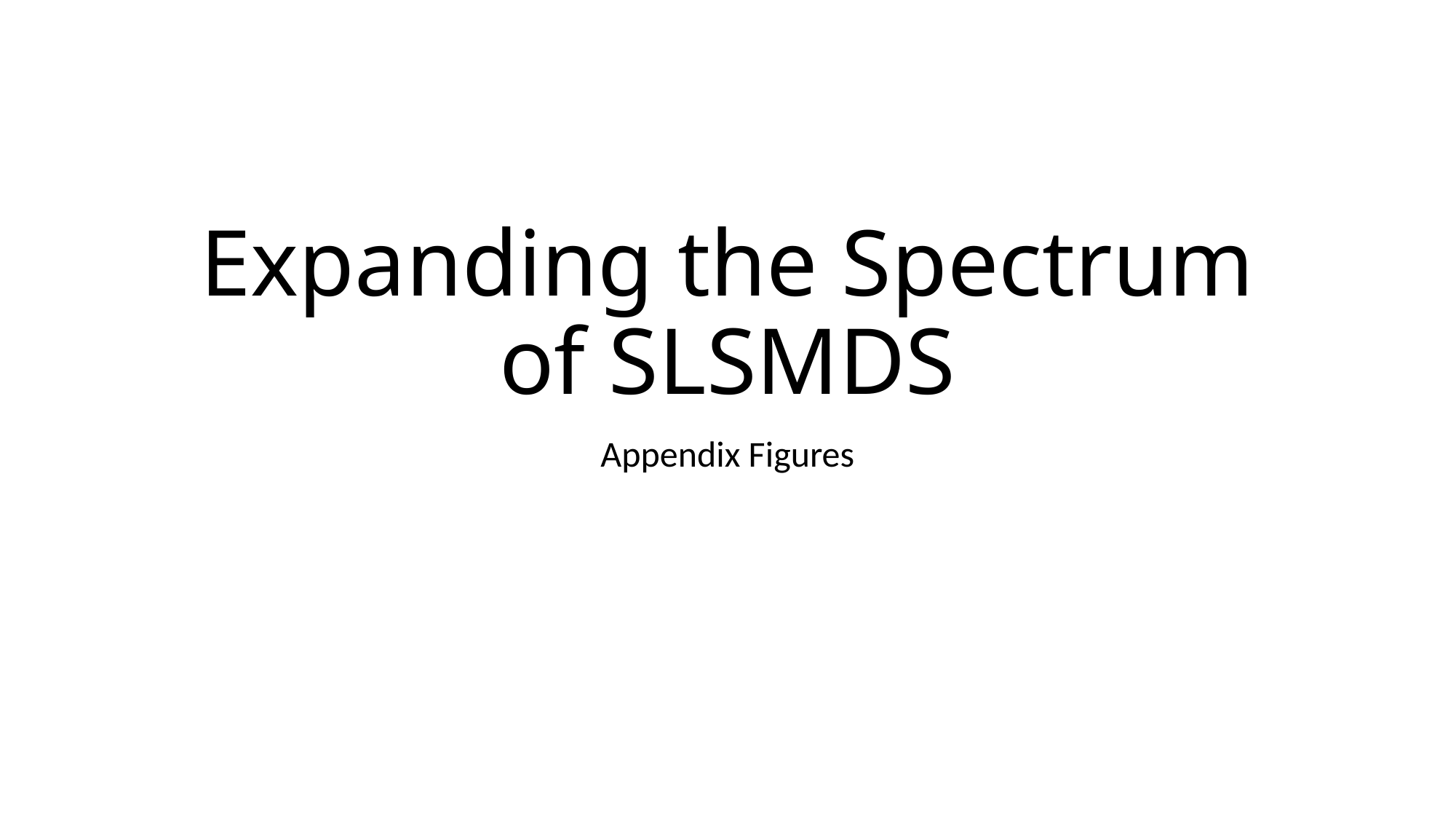

#### Expanding the Spectrum of SLSMDS
Appendix Figures

### Slide 2
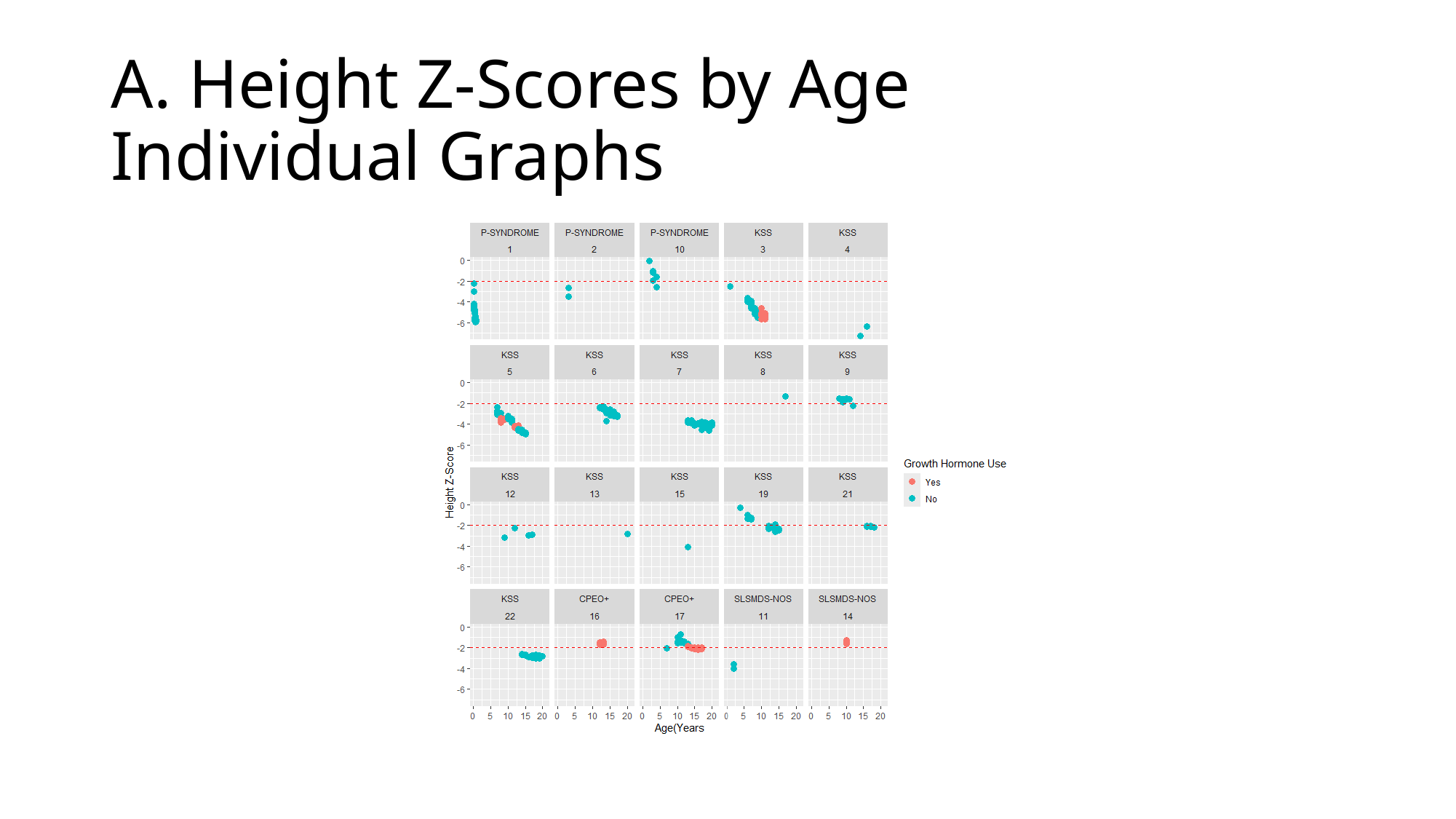

#### A. Height Z-Scores by Age Individual Graphs

### Slide 3
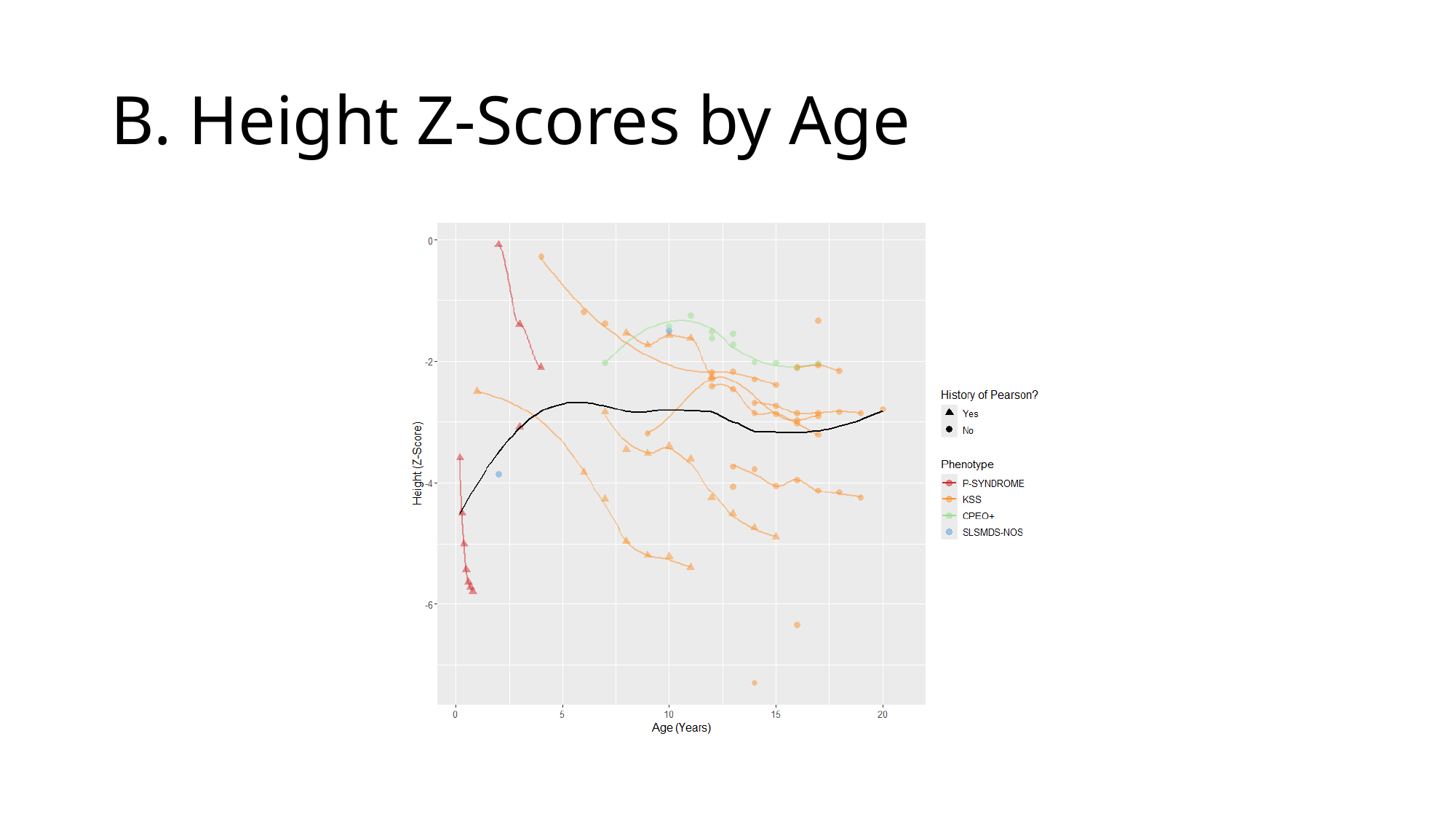

#### B. Height Z-Scores by Age

### Slide 4
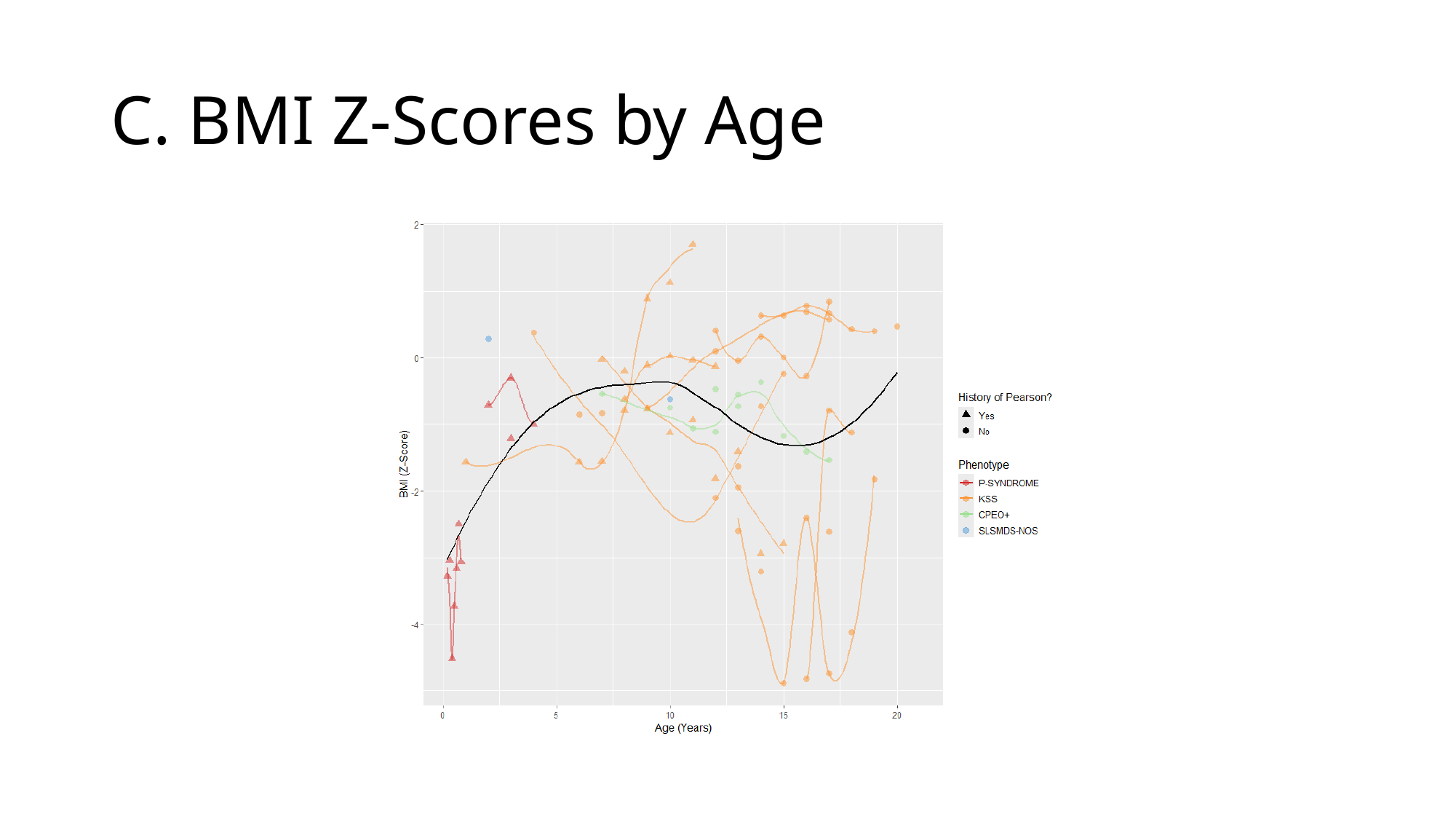

#### C. BMI Z-Scores by Age

### Slide 5
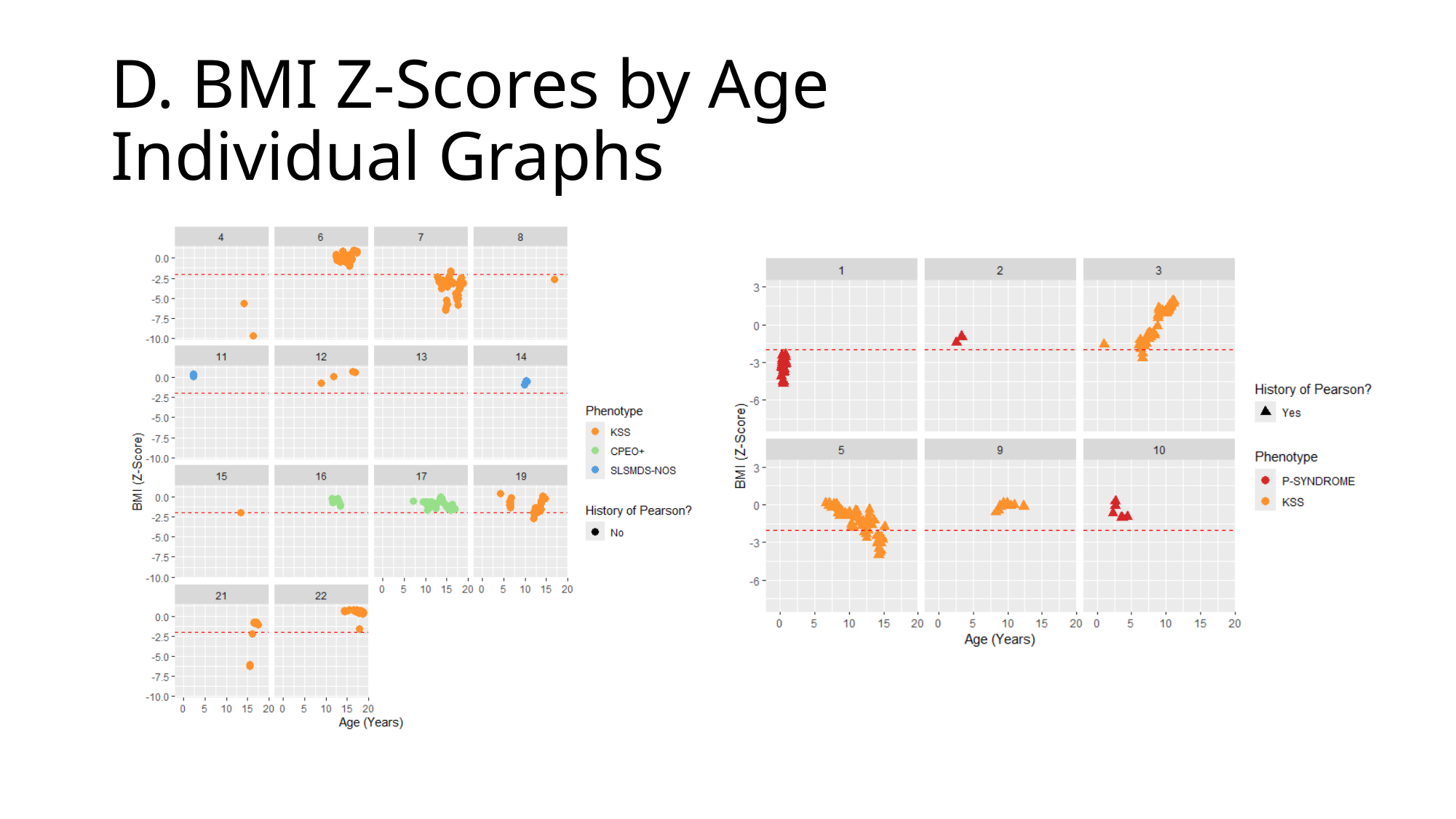

#### D. BMI Z-Scores by Age Individual Graphs

### Slide 6
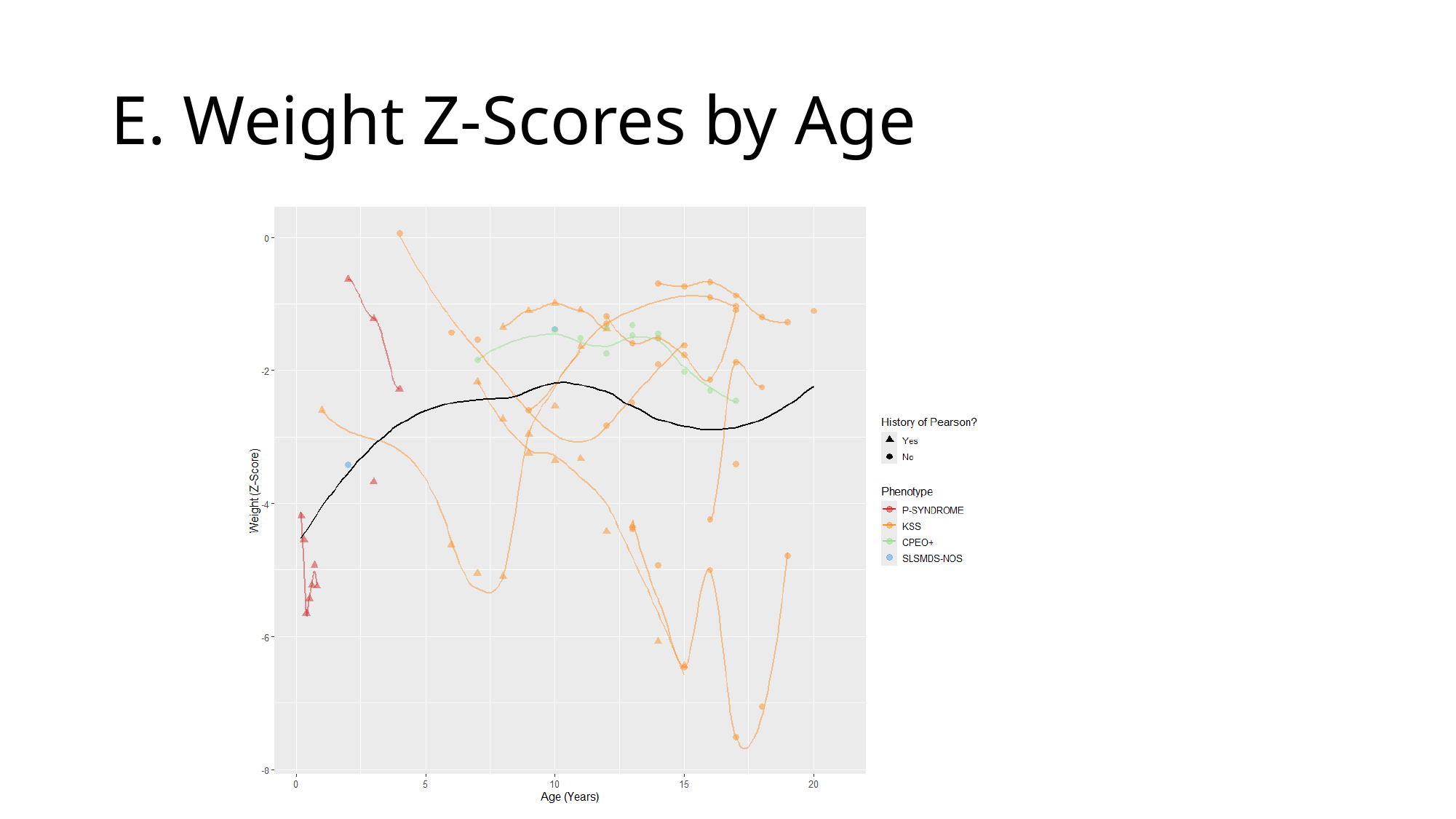

#### E. Weight Z-Scores by Age

### Slide 7
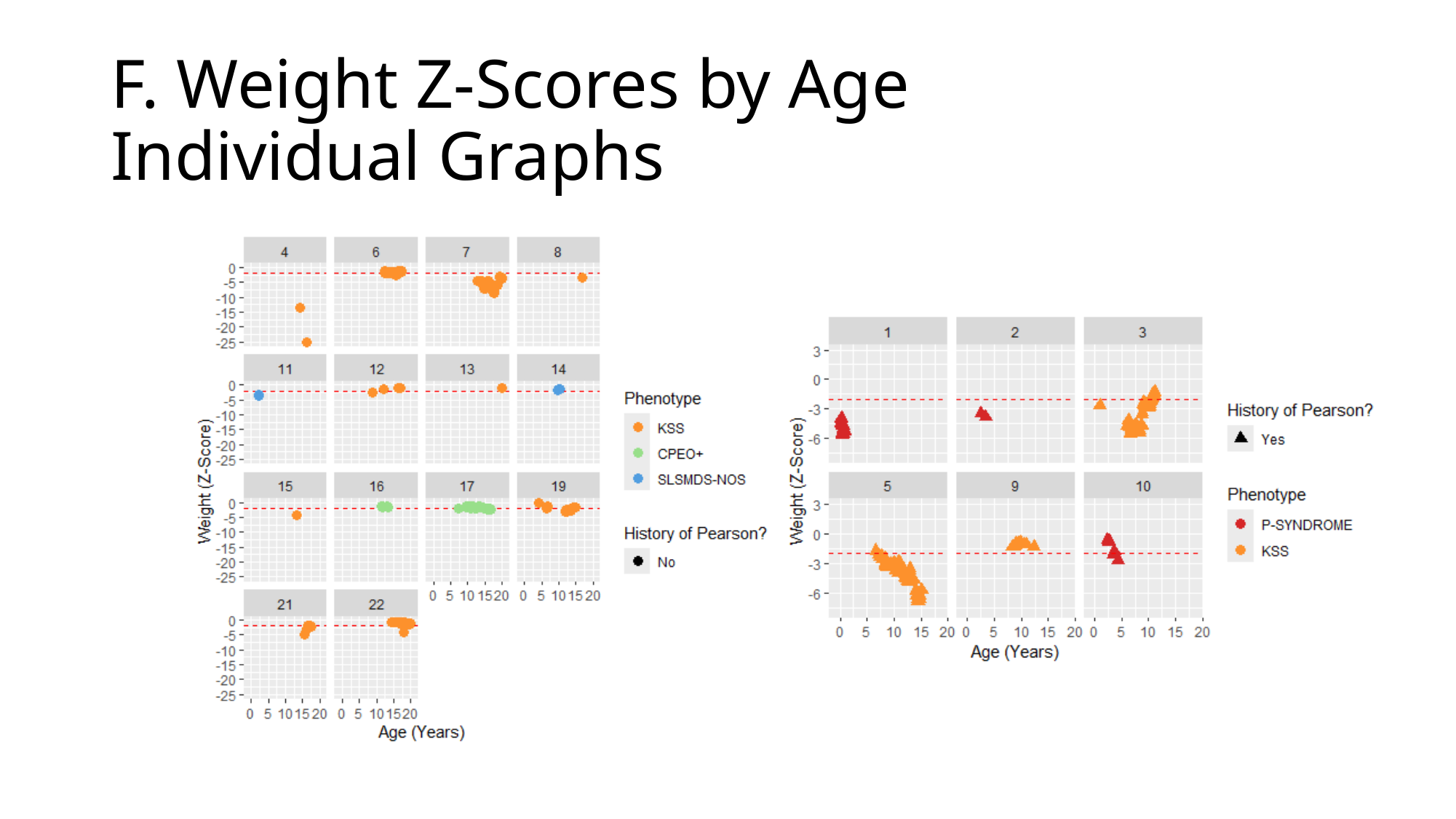

#### F. Weight Z-Scores by Age Individual Graphs

### Slide 8
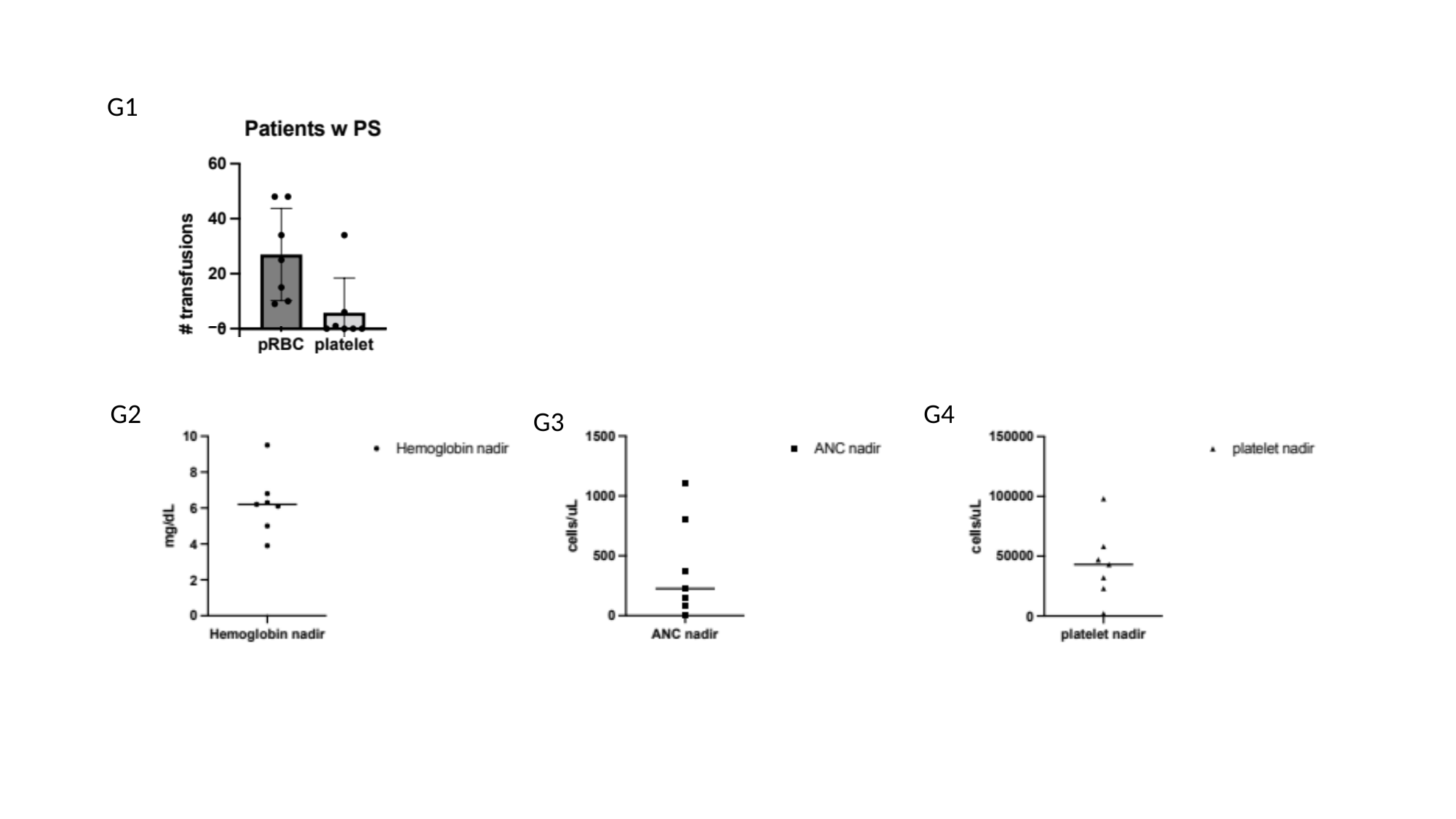

G1
G2
G4
G3

### Slide 9
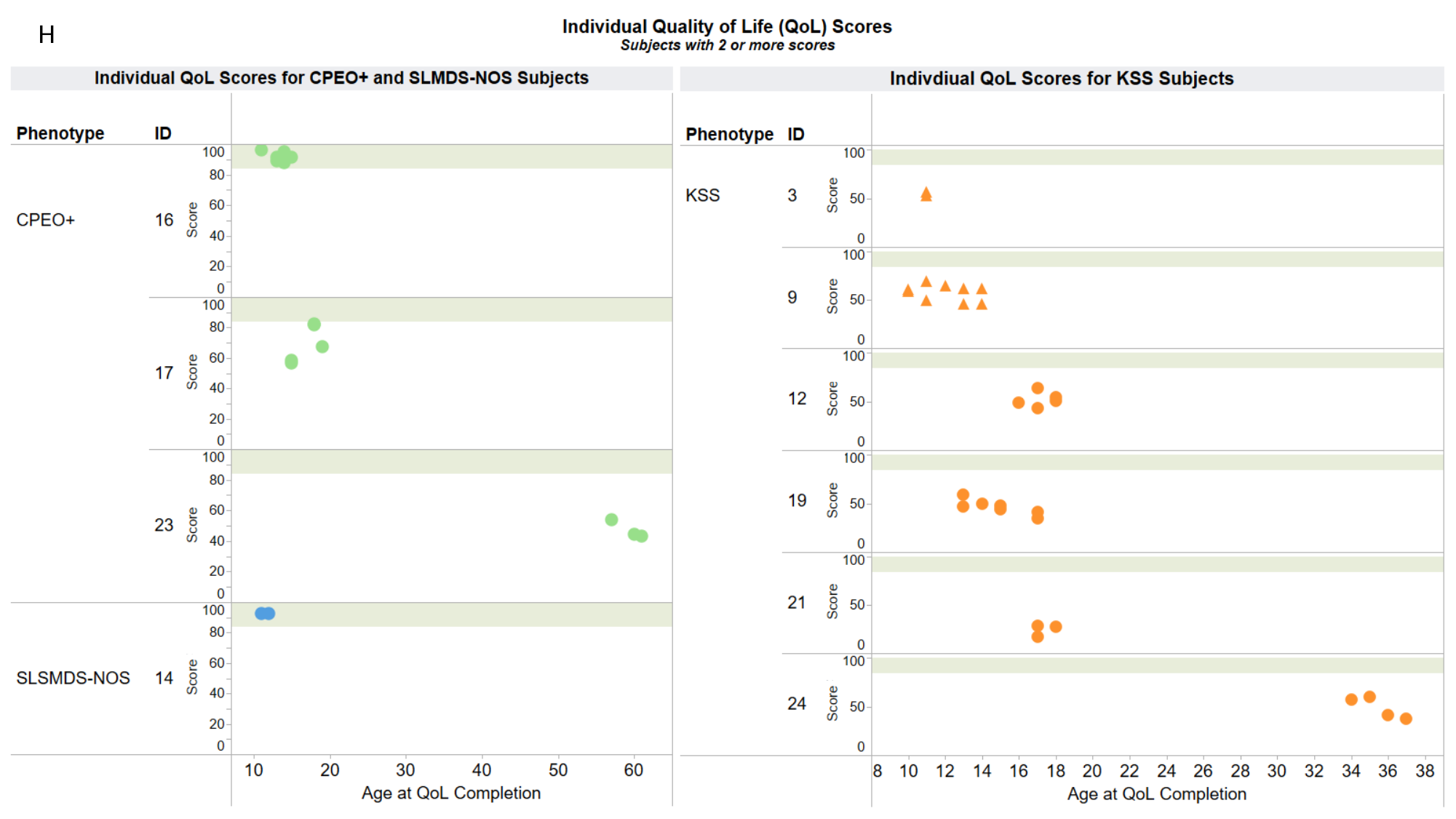

Appendix
H

### Slide 10
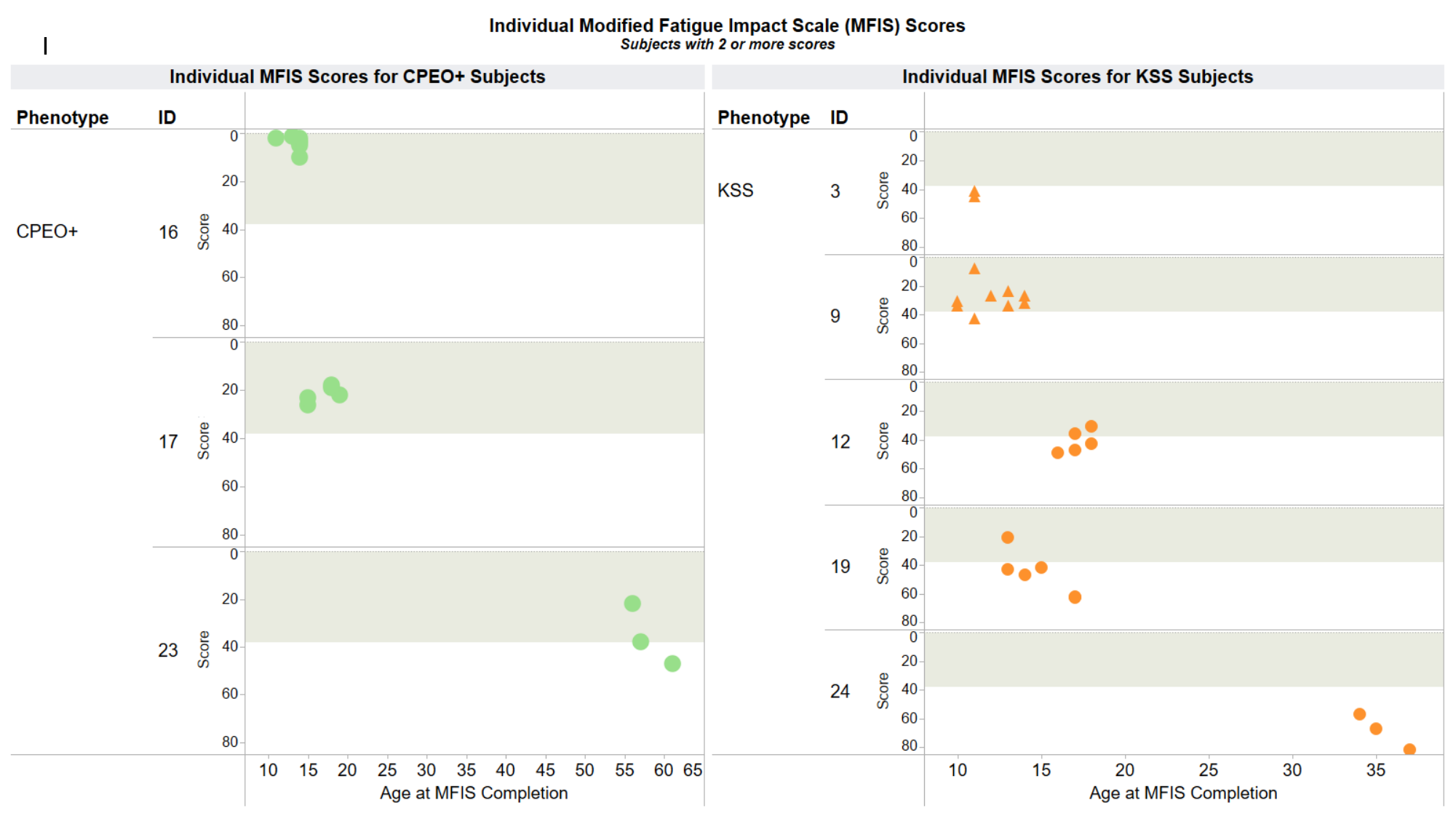

Appendix
I

### Slide 11
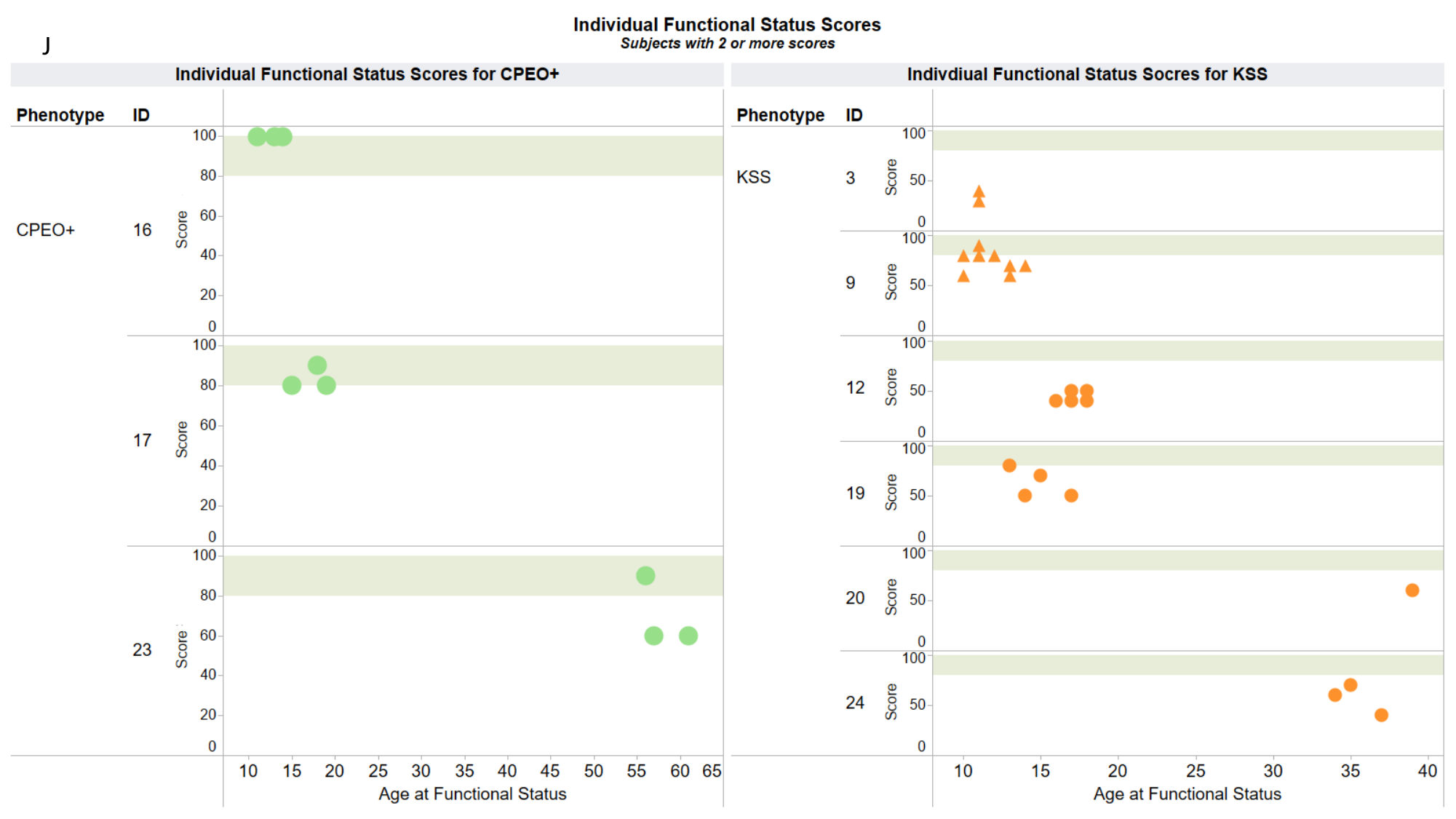

Appendix
J

### Slide 12
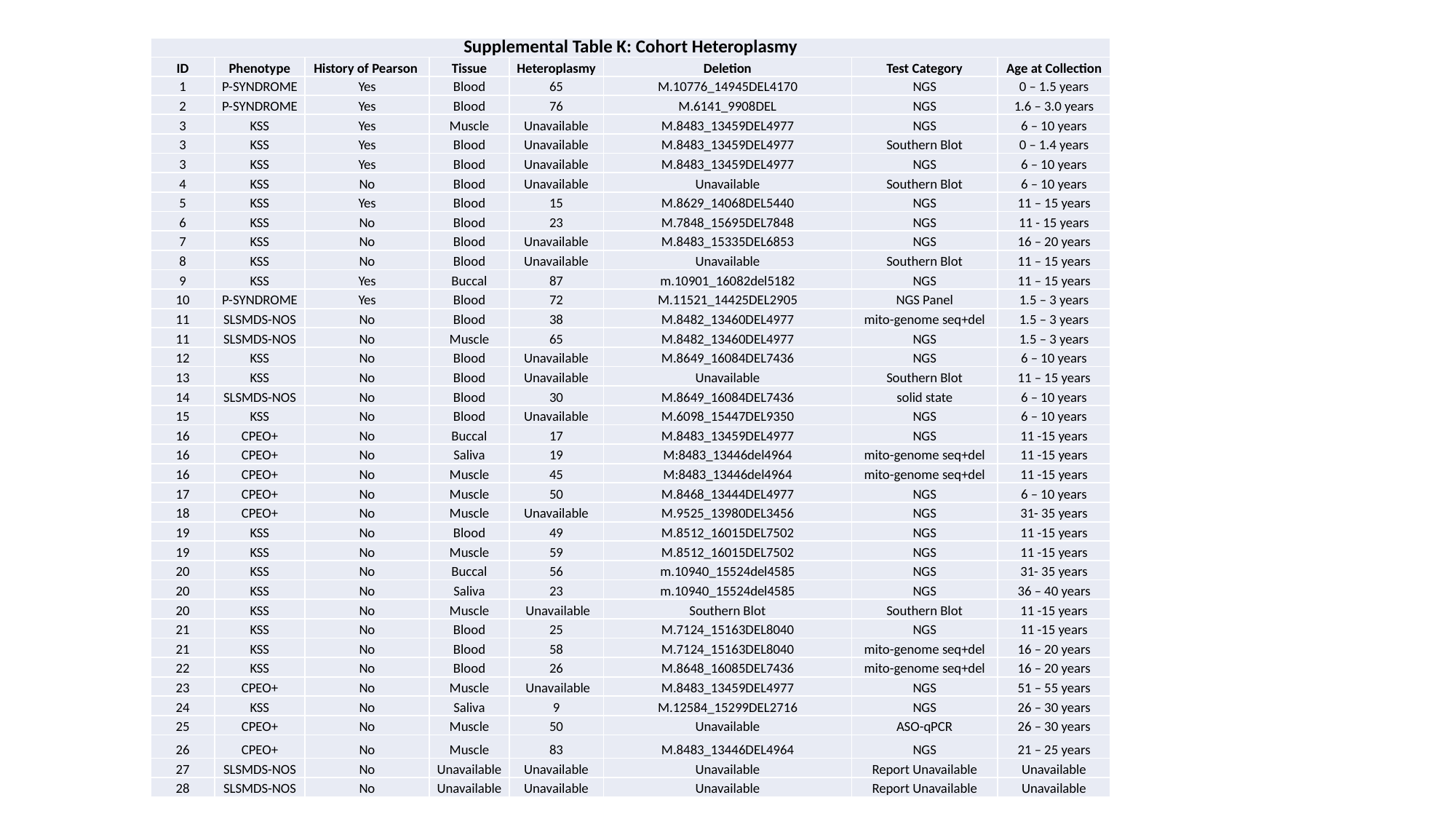

| Supplemental Table K: Cohort Heteroplasmy | | | | | | | |
| --- | --- | --- | --- | --- | --- | --- | --- |
| ID | Phenotype | History of Pearson | Tissue | Heteroplasmy | Deletion | Test Category | Age at Collection |
| 1 | P-SYNDROME | Yes | Blood | 65 | M.10776\_14945DEL4170 | NGS | 0 – 1.5 years |
| 2 | P-SYNDROME | Yes | Blood | 76 | M.6141\_9908DEL | NGS | 1.6 – 3.0 years |
| 3 | KSS | Yes | Muscle | Unavailable | M.8483\_13459DEL4977 | NGS | 6 – 10 years |
| 3 | KSS | Yes | Blood | Unavailable | M.8483\_13459DEL4977 | Southern Blot | 0 – 1.4 years |
| 3 | KSS | Yes | Blood | Unavailable | M.8483\_13459DEL4977 | NGS | 6 – 10 years |
| 4 | KSS | No | Blood | Unavailable | Unavailable | Southern Blot | 6 – 10 years |
| 5 | KSS | Yes | Blood | 15 | M.8629\_14068DEL5440 | NGS | 11 – 15 years |
| 6 | KSS | No | Blood | 23 | M.7848\_15695DEL7848 | NGS | 11 - 15 years |
| 7 | KSS | No | Blood | Unavailable | M.8483\_15335DEL6853 | NGS | 16 – 20 years |
| 8 | KSS | No | Blood | Unavailable | Unavailable | Southern Blot | 11 – 15 years |
| 9 | KSS | Yes | Buccal | 87 | m.10901\_16082del5182 | NGS | 11 – 15 years |
| 10 | P-SYNDROME | Yes | Blood | 72 | M.11521\_14425DEL2905 | NGS Panel | 1.5 – 3 years |
| 11 | SLSMDS-NOS | No | Blood | 38 | M.8482\_13460DEL4977 | mito-genome seq+del | 1.5 – 3 years |
| 11 | SLSMDS-NOS | No | Muscle | 65 | M.8482\_13460DEL4977 | NGS | 1.5 – 3 years |
| 12 | KSS | No | Blood | Unavailable | M.8649\_16084DEL7436 | NGS | 6 – 10 years |
| 13 | KSS | No | Blood | Unavailable | Unavailable | Southern Blot | 11 – 15 years |
| 14 | SLSMDS-NOS | No | Blood | 30 | M.8649\_16084DEL7436 | solid state | 6 – 10 years |
| 15 | KSS | No | Blood | Unavailable | M.6098\_15447DEL9350 | NGS | 6 – 10 years |
| 16 | CPEO+ | No | Buccal | 17 | M.8483\_13459DEL4977 | NGS | 11 -15 years |
| 16 | CPEO+ | No | Saliva | 19 | M:8483\_13446del4964 | mito-genome seq+del | 11 -15 years |
| 16 | CPEO+ | No | Muscle | 45 | M:8483\_13446del4964 | mito-genome seq+del | 11 -15 years |
| 17 | CPEO+ | No | Muscle | 50 | M.8468\_13444DEL4977 | NGS | 6 – 10 years |
| 18 | CPEO+ | No | Muscle | Unavailable | M.9525\_13980DEL3456 | NGS | 31- 35 years |
| 19 | KSS | No | Blood | 49 | M.8512\_16015DEL7502 | NGS | 11 -15 years |
| 19 | KSS | No | Muscle | 59 | M.8512\_16015DEL7502 | NGS | 11 -15 years |
| 20 | KSS | No | Buccal | 56 | m.10940\_15524del4585 | NGS | 31- 35 years |
| 20 | KSS | No | Saliva | 23 | m.10940\_15524del4585 | NGS | 36 – 40 years |
| 20 | KSS | No | Muscle | Unavailable | Southern Blot | Southern Blot | 11 -15 years |
| 21 | KSS | No | Blood | 25 | M.7124\_15163DEL8040 | NGS | 11 -15 years |
| 21 | KSS | No | Blood | 58 | M.7124\_15163DEL8040 | mito-genome seq+del | 16 – 20 years |
| 22 | KSS | No | Blood | 26 | M.8648\_16085DEL7436 | mito-genome seq+del | 16 – 20 years |
| 23 | CPEO+ | No | Muscle | Unavailable | M.8483\_13459DEL4977 | NGS | 51 – 55 years |
| 24 | KSS | No | Saliva | 9 | M.12584\_15299DEL2716 | NGS | 26 – 30 years |
| 25 | CPEO+ | No | Muscle | 50 | Unavailable | ASO-qPCR | 26 – 30 years |
| 26 | CPEO+ | No | Muscle | 83 | M.8483\_13446DEL4964 | NGS | 21 – 25 years |
| 27 | SLSMDS-NOS | No | Unavailable | Unavailable | Unavailable | Report Unavailable | Unavailable |
| 28 | SLSMDS-NOS | No | Unavailable | Unavailable | Unavailable | Report Unavailable | Unavailable |
